# The impact of SARS-CoV-2 on fatigue and quality of life in the early phase of the pandemic: prevalence of post COVID-19 condition among unvaccinated individuals in a Dutch population-based serosurveillance cohort

**DOI:** 10.1101/2024.03.19.24304303

**Authors:** Cheyenne C.E. van Hagen, Elizabeth N. Mutubuki, Siméon de Bruijn, Eric R.A. Vos, Gerco den Hartog, Fiona R.M. van der Klis, Cees C. van den Wijngaard, Hester E. de Melker, Albert Jan van Hoek

**Affiliations:** National Institute for Public Health and the Environment (RIVM), Center for Infectious Disease Control, Bilthoven, the Netherlands; National Institute for Public Health and the Environment (RIVM), Center for Health and Society, Bilthoven, the Netherlands; Laboratory of Medical Immunology, Radboud UMC, Nijmegen, The Netherlands

**Keywords:** Health-related quality of life, mental health, physical health, post-covid-19 condition, fatigue, SARS-CoV-2 infection, seroepidemiology

## Abstract

**Objectives:** We studied post-COVID-19 condition by investigating health-related quality of life and fatigue in the general Dutch population in the early phase of the pandemic, including symptomatic and asymptomatic infections among unvaccinated individuals.

**Methods:** (Still) unvaccinated participants aged ≥15 years were selected from the February 2021 round of the nationwide seroepidemiological PIENTER Corona cohort study. We assessed associations between the time since serologically-identified SARS-CoV-2 infection and four outcome measures: health utility (Short-Form 6 Dimensions), mental health and physical health (Short Form Health Survey 12) and fatigue (Checklist Individual Strength subscale fatigue). Per outcome, cutoff points were selected at each 5% increment (5-75%) along the cumulative distribution of those uninfected. At each cutoff, multivariable logistic regression models (score below cutoff yes/no) were fitted adjusted for infection history, age, sex, education level, comorbidities, and restriction intensity.

**Results:** At the cutoff of the lowest 15th percentile among uninfected, significant differences between uninfected (n=4,569) and infected ≤4 months ago (n=351) were observed for health utility (OR [95%CI]: 1.6 [1.2-2.2]), physical health (1.9 [1.5-2.5]) and fatigue (1.6 [1.3-2.1]), but not for mental health (1.2 [0.9-1.6]). There were no significant differences between uninfected and infected >4 months ago (n=327) for all outcomes at any cutoff of the cumulative distribution, with post-hoc analysis showing a power to detect prevalence differences as low as 7%.

**Conclusions:** In the first year of the pandemic, data from this Dutch population-based seroepidemiological cohort showed that unvaccinated individuals with a SARS-CoV-2 infection ≤4 months ago reported poorer health utility and physical health, and more severe fatigue compared to those uninfected. Interestingly, for those infected >4 months ago differences remained below the detection limit, suggesting a lower population prevalence of post-COVID-19 condition than currently found in literature for this period.

## Background

Identifying and measuring the overall impact of severe acute respiratory syndrome coronavirus 2 (SARS-CoV-2) on health-related quality of life (HRQoL) is crucial for informing public health strategies. During the pandemic, HRQoL of the general population was negatively affected as a result of both physical (e.g. infection or disease) and psychological (e.g. anxiety or social isolation) factors (1–3). With some groups more at risk of (severe) SARS-CoV-2 infection, different groups were disproportionately impacted (4). Clinical presentations of SARS-CoV-2 can range from no or mild symptoms to critical illness and death. In a proportion of patients, symptoms develop or remain after the acute phase resulting in long-term complaints, a condition referred to as Long COVID or Post-COVID-19 Condition (PCC) (5–8). PCC is defined by the World Health Organization as “the continuation or development of new symptoms 3 months after the initial SARS-CoV-2 infection, with these symptoms lasting for at least 2 months with no other explanation” (9). Symptoms of PCC include shortness of breath and cognitive dysfunction, with the most common symptom being fatigue (9, 10). Due to the long duration and/or detrimental impact of these symptoms, PCC may form a significant proportion of the overall disease burden of SARS-CoV-2 (11). Therefore, the impact of PCC on the population in terms of HRQoL is important to investigate.

The burden of PCC in the population depends on SARS-CoV-2 infection rates and the proportion of individuals developing PCC after infection. Although the prevalence of PCC has been studied in prospective cohorts and in cross-sectional population samples, where patients are identified by a positive SARS-CoV-2 PCR or lateral flow test (12, 13), discrepancies exist in PCC prevalence estimates (ranging from 9% to 81%) (10). These discrepencies predominantly arise due to variations in definitions of PCC, the population under investigation, the identification of (re-)infection/COVID-19, or other factors such as SARS-CoV-2 variant, time since infection or COVID-19 vaccination. Vaccination and previous infection contribute to immunological heterogeneity in current populations, making it difficult to establish PCC prevalence after more recent SARS-CoV-2 variants (14). To avoid vaccination effects on PCC development, studying early-pandemic SARS-CoV-2 infections, before the rollout of vaccination programs, is insightful and could function as a benchmark for more recent SARS-CoV-2 variants amidst mostly vaccinated populations. In addition, studies that rely on positive SARS-CoV-2 testing are prone to selection bias due to testing behavior and asymptomatic infections. Therefore, to establish the true prevalence of PCC and its impact on HRQoL on a population level, population-based serological studies are required because such studies provide a more comprehensive and unbiased assessment of the infection rates in the general population (15).

Although a population-based and/or serological approach was previously used to assess PCC in some studies (16–19), these often involved a more heterogenous immunological landscape, included only children or focused on a variety of PCC-related symptoms rather than looking at the HRQoL. To our knowledge, there are no population-based serological studies investigating HRQoL after SARS-CoV-2 infection in the pre-vaccination era. Hence, using Dutch population-based serosurveillance data from the first year of the COVID-19 pandemic, the aim of this study was to assess the relationship between the time since wild-type SARS-CoV-2 infection and HRQoL and fatigue as endpoints linked to PCC among unvaccinated individuals.

## Materials and methods

### Study design and population

The PIENTER Corona (PICO) study is a prospective population-based cohort study aimed at monitoring humoral immunity against SARS-CoV-2 in the Dutch population. The PICO study started in April 2020 (PICO1) when 3,244 participants aged 2-92 years were enrolled (20). These participants were sampled from the PIENTER3 serosurvey, an existing cohort established in 2016/2017 (21), for which participants had been randomly selected (using age strata) from the Dutch population registry. In the second round, in June 2020 (PICO2), the study sample was supplemented with an additional sample of 4,606 randomly-selected participants from the Dutch population registry in order to enhance national coverage and overall power (combined response rate, 21.4%) (22). Subsequent rounds were conducted in September 2020 (PICO3) and February 2021 (PICO4) using the same study sample as in PICO2 with exception to dropouts and non-responders (n=1,320) (Figure 1).

**Figure 1.**
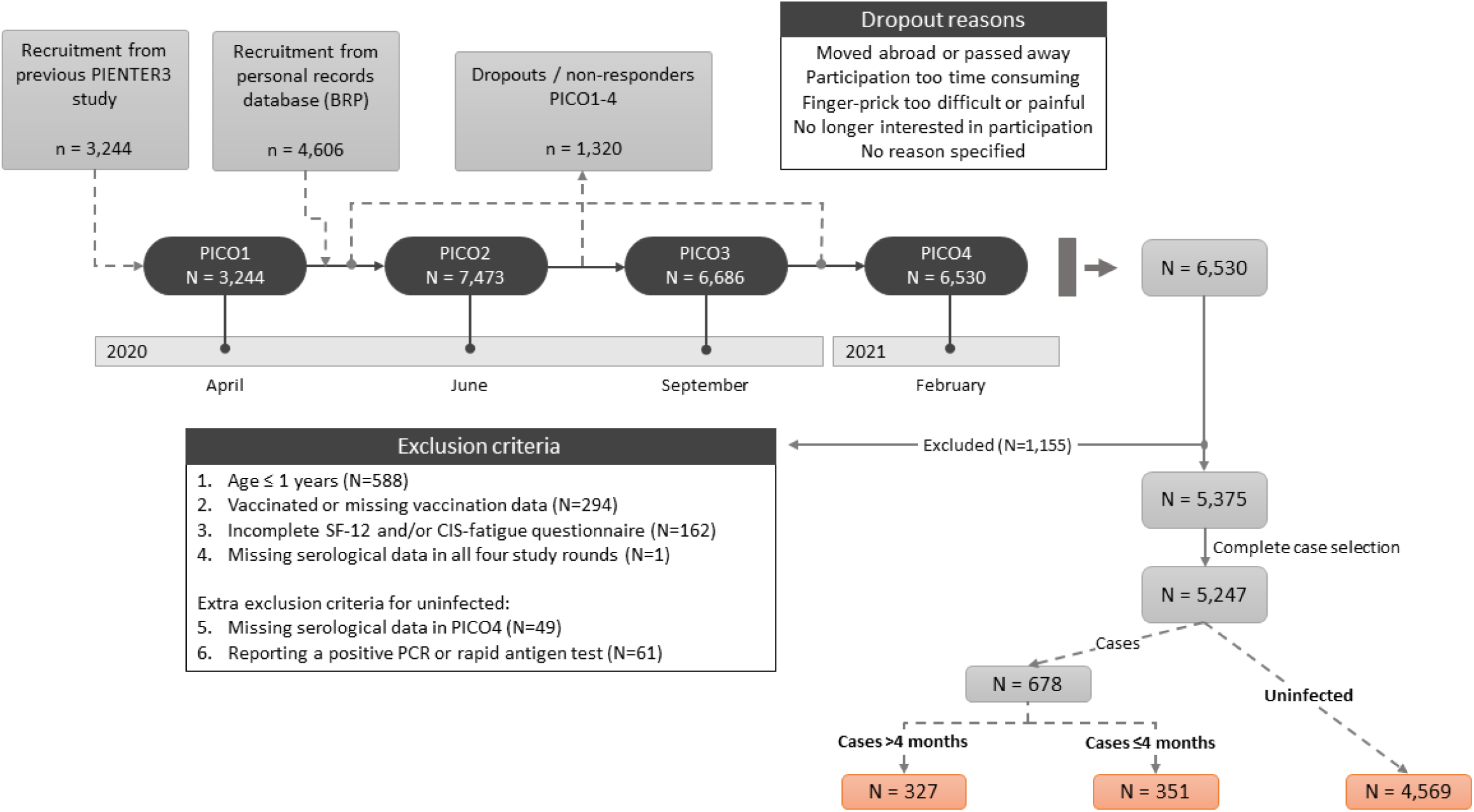
Flowchart of participant recruitment in PICO1-4

In every study round, participants were asked to return a finger-prick blood sample in a microtainer by mail and complete a questionnaire (online or on paper). The questionnaire collected information on demographics, experienced COVID-19 like symptoms (i.e., respiratory, gastro-intestinal, systemic, etc.), SARS-CoV-2 testing, COVID-19 vaccination, clinical risk factors for disease, contact patterns, HRQoL and fatigue (only from PICO4 onwards for those 15 years and older). Blood samples were tested for immunoglobulin G (IgG) antibodies against SARS-CoV-2 Spike-S1 and Nucleoprotein (N) proteins using a previously described validated fluorescent bead-based multiplex-immunoassay (23). Briefly, serum samples were centrifuged and stored at −20 degrees Celsius before analysis. The analysis process included dilution and incubation of samples with S1-(Sino Biological, 40591-V08H) and N-(Sino Biological, 40588-V08B) coupled beads in SM01-buffer (Surmodics, USA) supplemented with 2% FCS and shaking of samples for 45 minutes in the dark at room temperature. Subsequently, plates underwent three washes with PBS and were incubated for 30 minutes with PE-conjugated goat anti-human IgG (Jackson ImmunoResearch, 109-116-098) and washed again. Finally, samples were acquired on a Luminex LX200 or FlexMap3D. A 5-parameter logistic fit was used to interpolate antibody concentrations from pooled sera calibrated against the World Health Organization standard (NIBSC, 20/136). The cut-off for seropositivity was set at a Spike-S1-specific IgG antibody concentration of 10.08 binding antibody units per milliliter (BAU/mL) and a N-specific IgG antibody concentration of 14.3 BAU/mL, in accordance with international standards (22–25). This yielded a specificity and a sensitivity of 99.9% and 94.3% respectively for anti-S1 and 98.0% and 86.0% for anti-N (22, 24–26).

In this study we included all unvaccinated PICO4 participants with questionnaire data on HRQoL and fatigue and at least one blood sample in PICO1-4. COVID-19 vaccination status was based on the Dutch COVID-19 vaccination registry (CIMS) (27) and complemented with self-reported questionnaire data if CIMS was unavailable. The PICO study protocol was approved by the Medical Ethics Committee MEC-U, the Netherlands (Clinical Trial Registration NTR8473). All participants provided written informed consent.

### Time since SARS-CoV-2 infection

To assess SARS-CoV-2 infection, we distinguished between participants with serological proof of previous SARS-CoV-2 infection (seropositives/cases) and those without (seronegatives/uninfected). Proof of infection was defined as the presence of serum immunoglobulin G (IgG) antibodies against the SARS-CoV-2 Spike-S1 antigen in rounds PICO1-4. Participants who were Spike-S1 seropositive at least once were considered a case, irrespective of negative PCR/rapid antigen tests or serostatus in subsequent rounds. To assess time since SARS-CoV-2 infection, cases were further classified according to time since seroconversion; we distinguished participants that seroconverted in PICO1-3, i.e., before or in September 2020 (cases >4 months) and participants that seroconverted in PICO4, i.e., after September 2020 (cases ≤4 months). The uninfected group consisted of all participants who had not been Spike-S1 seropositive through PICO1-4. Uninfected participants who missed serological data in PICO4 or self-reported a positive PCR or rapid antigen test in one of the study rounds were excluded. Thus, three SARS-CoV-2 serostatus groups were ultimately identified: uninfected, cases >4 month and cases ≤4 months.

### Outcome measures

#### HRQoL: Short Form Health Survey 12 (SF-12) and Short-Form Six Dimensions (SF-6D)

HRQoL was assessed in terms of mental health, physical health and health utility. To measure these, the Dutch translation of the SF-12 version 1 was used. The SF-12 consists of 12 questions from eight health dimensions that can be summarized into a physical health score (physical component summary; PCS) and a mental health score (mental component summary; MCS). The PCS comprises the following health dimensions; physical functioning, role limitations due to physical health problems, bodily pain and general health, whereas the MCS consists of vitality (energy/fatigue), social functioning, role limitations due to emotional problems and mental health (psychological distress and psychological well-being). The summarized scores were weighted using orthogonal regression coefficients from the general Dutch population (28). PCS and MCS scores range from 0 (lowest health) to 100 (highest health) (29). To convert results of the SF-12 into health utility scores the SF-6D was used (30). The SF-6D uses seven items from the SF-12 and consists of six dimensions: physical functioning, role limitations, social functioning, pain, mental health and vitality (30). SF-6D scores range from 0.3 to 1, with higher scores indicating better health (30, 31). In the online questionnaire, participants received the SF-12 version 1 UK, while the paper-based questionnaire contained the SF-12 version 1 US. In the SF-12 version 1 UK, item 12 (social functioning) has a six-scale answer instead of a five-scale answer that is used in the SF-12 version 1 US. Participants who selected the additional answer option (i.e. option 3 of the six-scale answer) in the SF-12 version 1 UK were equally randomized to a category below and a category above in order to analyze all questionnaires in the SF-12 version 1 US format.

#### Fatigue: Checklist Individual Strength (CIS)

Fatigue severity was assessed using the fatigue subscale of the Checklist Individual Strength (CIS-fatigue). The CIS-fatigue is an 8-item questionnaire validated to assess chronic fatigue (32, 33). Each item is scored on a 7-point Likert scale. Scores can range from 8 to 56, with fatigue severity increasing with higher scores (34).

### Statistical analysis

Participant demographic (age, sex and educational level) and health characteristics (number of comorbidities, health utility, physical health, mental health and fatigue) were presented using proportions (%) and frequencies (n). Information collected in PICO4 were used, except for educational level and number of comorbidities which were determined at baseline.

In the analyses we looked at four different outcomes: health utility (SF-6D), mental health (MCS), physical health (PCS), and fatigue (CIS). To enable comparison between the four outcome variables with different scales and score ranges, we opted for a uniform approach. In this approach, we separately plotted the cumulative distribution of each outcome for all three SARS-CoV-2 serostatus groups to visualize differences between these groups over the entire range of the outcome. Since higher CIS scores indicate worse outcomes, the cumulative distribution of the CIS scale was inverted for comparability. The cumulative distribution of individuals in the uninfected group guided the selection of cut-off points. Cut-off points were determined at each 5% increment along the cumulative distribution of the uninfected group (5% to 75%), and at each cut-off individuals were assigned 0 (score above the cut-off) or 1 (score on or below the cut-off). For each of the cut-off points multivariable logistic regression models were fitted separately, per outcome. The following confounders were added to the multivariable logistic regression models: age group (18-35/36-65/66+ years), sex (male/female), educational level (low/intermediate/high), number of comorbidities (none/one/two or more) and Stringency Index (75/79). The Stringency Index measures the intensity of restrictions in the Netherlands on a scale of 0 to 100 (strictest) as quantified by the COVID-19 Stringency Index of the Oxford Coronavirus Government Response Tracker (OxCGRT) (35, 36). The Stringency Index was dichotomized into score 75 and 79 because the scores remained steady during the inclusion period (78.7 from 11 February to 2 March 2021 and 75.0 from 3 March to 6 April 2021). Complete case data on all variables were used for the analyses. Adjusted odds ratios (OR) and 95% confidence intervals (CI) were provided and a p-value of p<0.05 was considered statistically significant.

Post-hoc analyses were conducted to explore the power to detect a minimal difference in prevalence between cases >4 months and those uninfected given the observed cases in our study. With those uninfected as reference group, an iterative process was performed in which per outcome, the prevalence of cases >4 months meeting the cut-off (assigned a 1) was increased until the CI of the OR was above 1. In this process the cases >4 months were included at random. To account for stochasticity, an average of ten independent samples was taken for each prevalence point, for each cut-off point.

Data was cleaned in SAS (94 M7 English) and analyzed in R (version 4.1.0).

## Results

### Demographic and clinical characteristics of the participants

From a total of 6,530 participants, 1,155 participants did not meet inclusion criteria of which 294 (4.5%) were excluded because of being vaccinated or missing vaccination data (Figure 1). Of 5,375 eligible participants (supplementary file – Table 1), complete case analysis was conducted with 5,247 participants of which 4,569 were uninfected, 327 cases >4 months and 351 cases ≤4 months (Table 1), thus excluding 2% of participants (missings). All groups had a higher proportion of females compared to males (55-58%). The majority of the participants in all groups were aged between 36-65 years (48-52%) and had a high or intermediate educational level (79-82%). Among cases ≤4 months there were fewer participants with comorbidities (27%) compared to those uninfected and cases >4 months (respectively 34% and 35%). In all groups, mean SF-6D scores were 0.8. Mean MCS (46–48) and PCS scores (53–55) were also similar between groups. Mean CIS scores of those uninfected and cases >4 months were comparable (22), but those of cases ≤4 months deviated (25).

**Table 1.** Participant demographic and health characteristics*.

|  | UNINFECTED | CASES >4 MONTHS | CASES ≤4 MONTHS |
| --- | --- | --- | --- |
| **N** | 4,569 | 327 | 351 |
| **SEX = FEMALE % (N)** | 55.5 (2,534) | 55.4 (181) | 57.8 (203) |
| **AGE GROUPS % (N)** |  |  |  |
| 15-35 | 21.7 (990) | 28.7 (94) | 32.2 (113) |
| 36-65 | 52.0 (2,375) | 47.7 (156) | 50.4 (177) |
| 66+ | 26.4 (1,204) | 23.5 (77) | 17.4 (61) |
| **EDUCATIONAL LEVEL<sup>A</sup> % (N)** |  |  |  |
| HIGH | 47.7 (2,178) | 43.7 (143) | 38.7 (136) |
| INTERMEDIATE | 31.8 (1,455) | 37.9 (124) | 39.9 (140) |
| LOW | 20.5 (936) | 18.3 (60) | 21.4 (75) |
| **NUMBER OF COMORBIDITIES<sup>B</sup> % (N)** |  |  |  |
| NONE | 65.9 (3,010) | 64.8 (212) | 73.2 (257) |
| ONE | 24.6 (1,126) | 27.2 (89) | 19.9 (70) |
| TWO OR MORE | 9.5 (433) | 8.0 (26) | 6.8 (24) |
| **HEALTH UTILITY (SF-6D) MEAN (SD)** | 0.82 (0.11) | 0.81 (0.11) | 0.79 (0.13) |
| **MENTAL HEALTH (MCS) MEAN (SD)** | 47.83 (10.26) | 47.16 (11.13) | 46.36 (10.87) |
| **PHYSICAL HEALTH (PCS) MEAN (SD)** | 54.53 (7.54) | 54.93 (6.86) | 53.18 (8.74) |
| **FATIGUE (CIS) MEAN (SD)** | 21.67 (11.67) | 21.73 (11.44) | 24.77 (12.96) |

\* All characteristics were determined in PICO4, except for educational level and comorbidities (determined at baseline).
<sup>A</sup> Educational level was classified as low (no education or primary education), intermediate (secondary school or vocational training), or high (bachelor's degree, university).
<sup>B</sup> Included comorbidities are asthma, cancer, cardiovascular disease, diabetes (type 1 & 2), immunodeficiency, pulmonary disease and renal disease.

### Cumulative distribution

For health utility, the cumulative distribution of SF-6D scores showed that cases ≤4 months had a higher proportion of individuals with a lower score, compared to those uninfected and cases >4 months (Figure 2, upper panel); among those uninfected and cases >4 months respectively 16% and 17% had an SF-6D score below 0.66, whereas for cases ≤4 months this was 22% (supplementary file – Table 2). Looking at mental health, cases ≤4 months had a higher proportion of individuals with lower MCS scores compared to those uninfected, which was also observed for cases >4 months across a smaller part of the cumulative distribution (Figure 3, upper panel); 15% of those uninfected scored below MCS 37, whereas for cases >4 months and cases ≤4 months this was respectively 18% and 19% (supplementary file – Table 3). For physical health, cases ≤4 months also had a higher proportion of individuals with lower PCS scores compared to those uninfected and cases >4 months (Figure 4, upper panel); among cases ≤4 months 22% had a PCS score below 48, whereas for those uninfected and cases >4 months this was 15% and 13% respectively (supplementary file – Table 4). For fatigue, cases ≤4 months had a higher proportion of individuals with higher CIS scores throughout most of the cumulative distribution (Figure 5, upper panel); 15% of those uninfected and cases >4 months scored above 35, whereas for cases ≤4 months this was 23% (supplementary file – Table 5).

**Figure 2.**
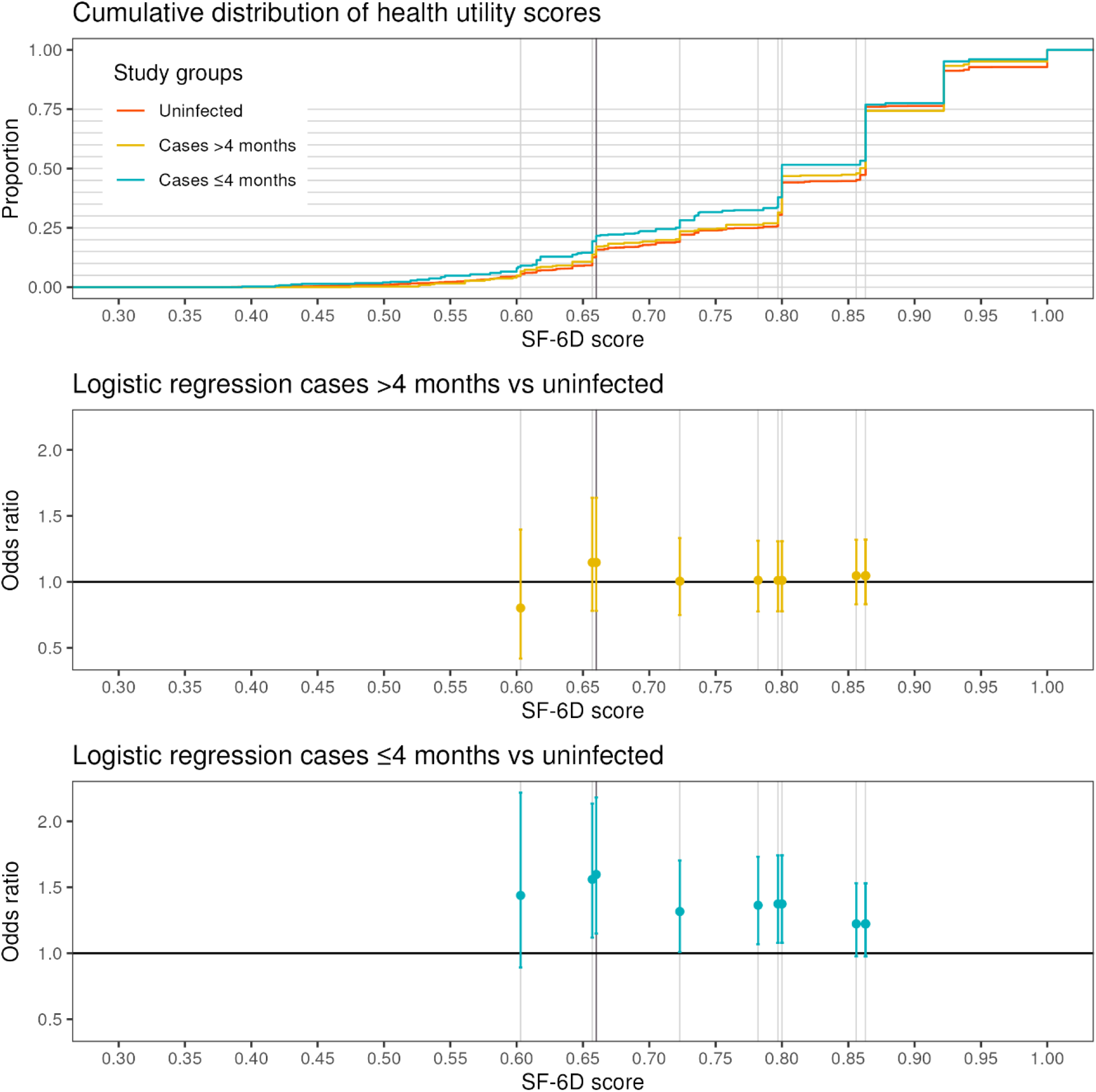
Health utility – cumulative distribution and logistic regression analyses^*^ ^*^Each 5% increment (5-75%) along the cumulative distribution of the control group is marked with a grey bar. The 15% point of the distribution is marked with a dark grey bar.

**Figure 3.**
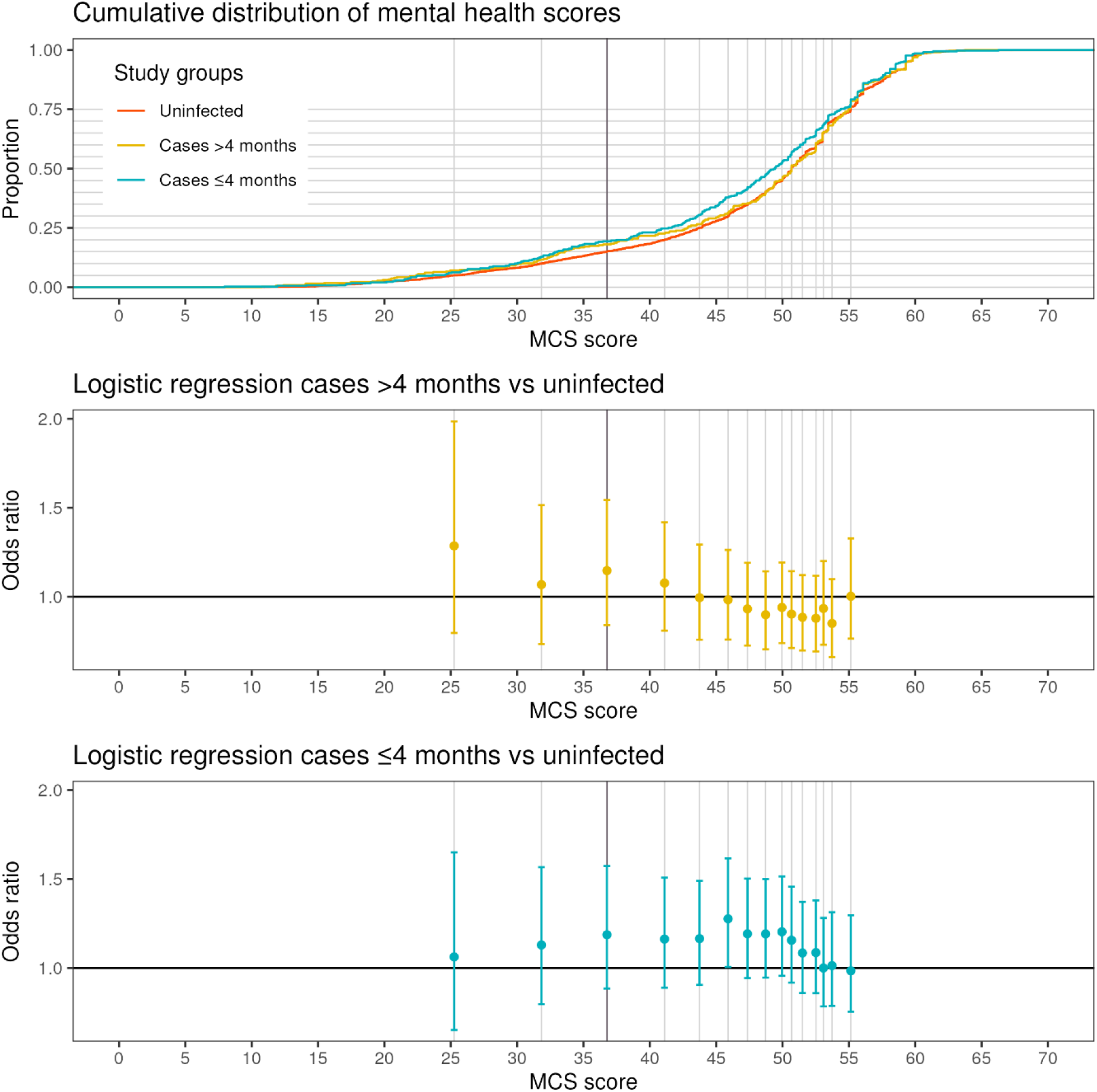
Mental health – cumulative distribution and logistic regression analyses^*^ ^*^Each 5% increment (5-75%) along the cumulative distribution of the control group is marked with a grey bar. The 15% point of the distribution is marked with a dark grey bar.

**Figure 4.**
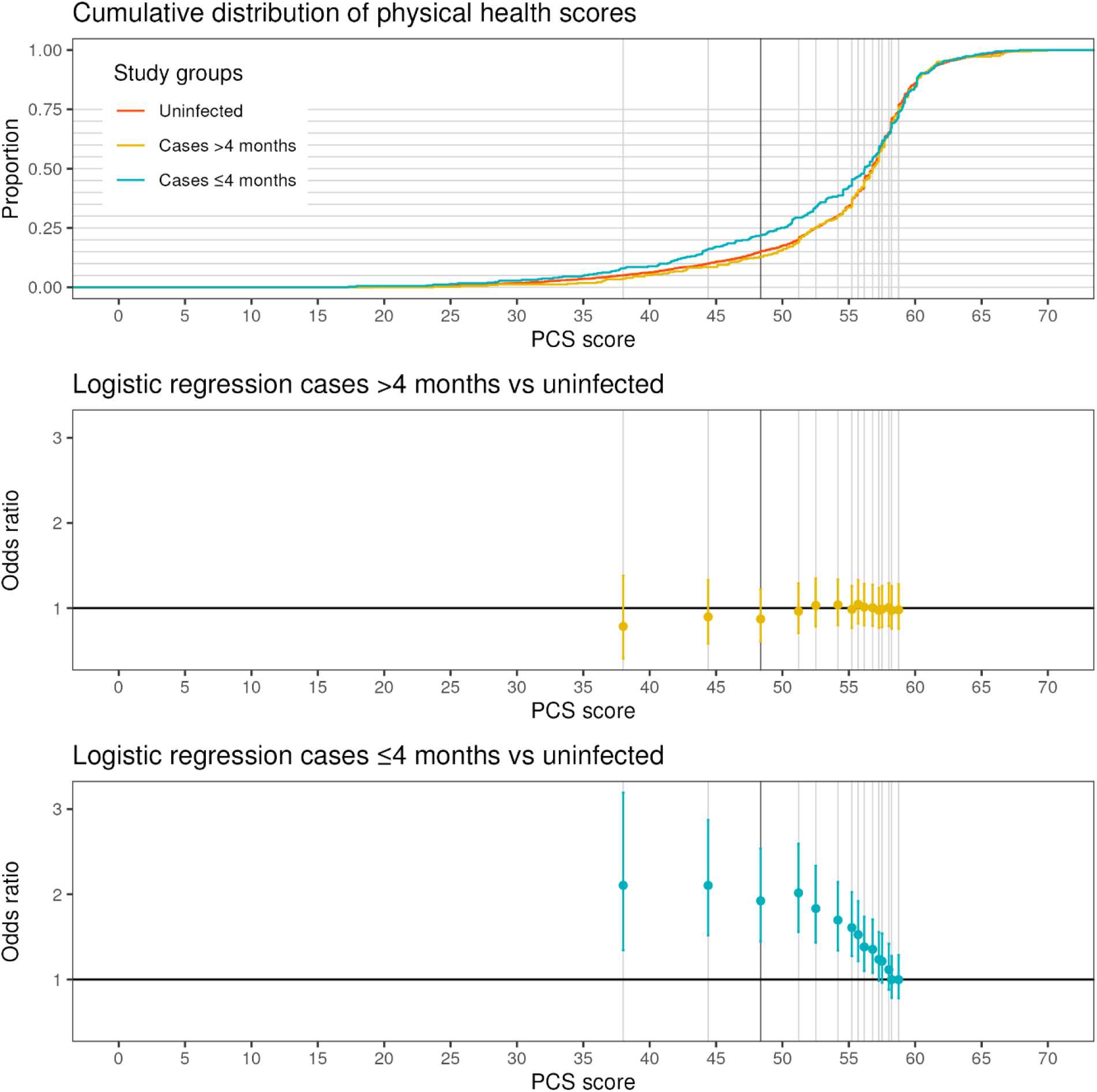
Physical health – cumulative distribution and logistic regression analyses^*^ ^*^Each 5% increment (5-75%) along the cumulative distribution of the control group is marked with a grey bar. The 15% point of the distribution is marked with a dark grey bar.

**Figure 5.**
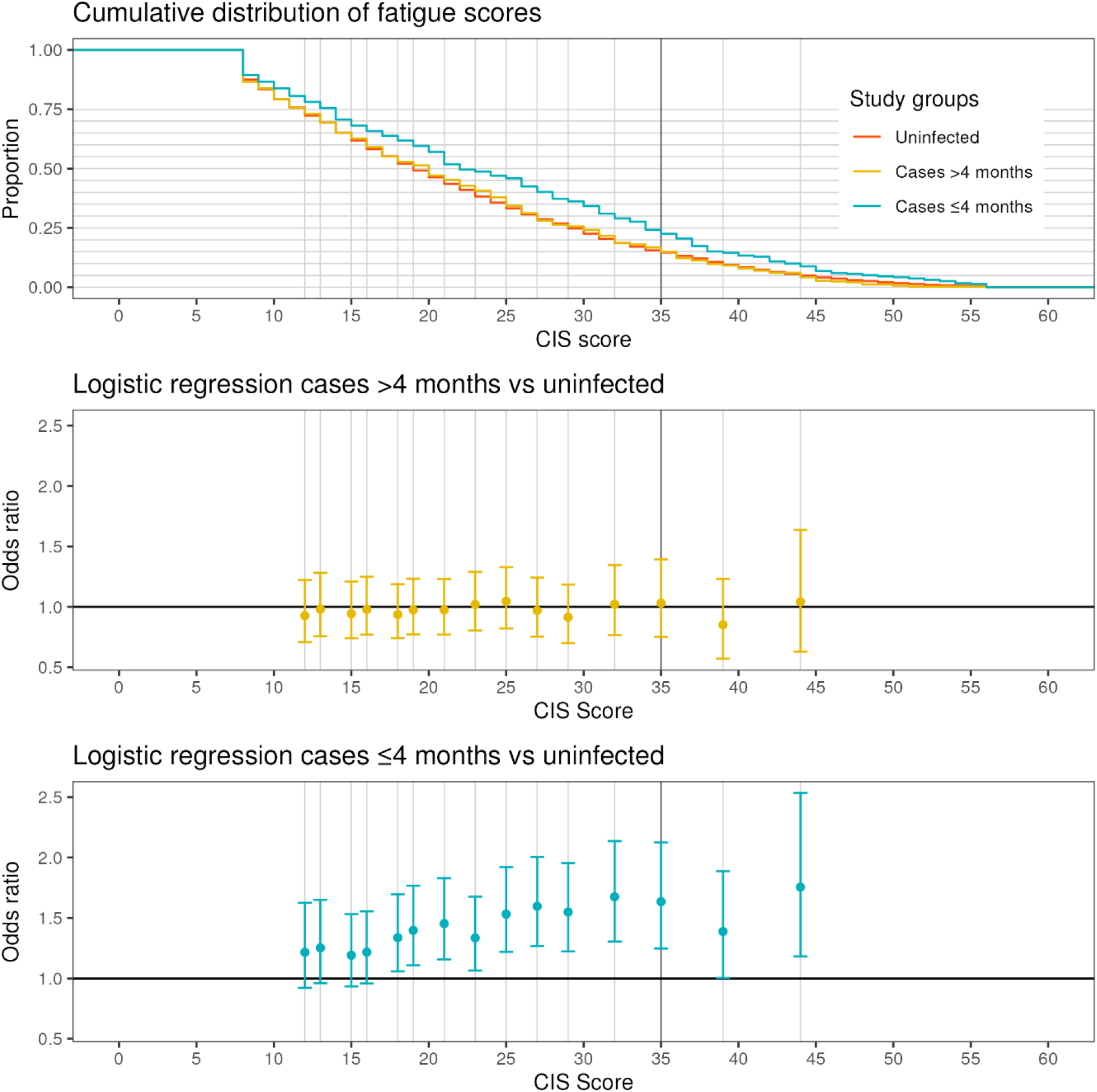
Fatigue – cumulative distribution and logistic regression analyses^*^ ^*^Each 5% increment (5-75%) along the cumulative distribution of the control group is marked with a grey bar. The 15% point of the distribution is marked with a dark grey bar.

### Multivariable logistic regression

The results of the logistic regression analyses showed that statistically significant differences between the groups differed based on the cut-off point of the outcome used. The 15% cut-off point corresponds to an SF-6D score of 0.66, an MCS score of 36.77, a PCS score of 48.36 and a CIS score of 35 (supplementary file – Table 2-5). This cut-off represents clinically relevant severity for the fatigue (CIS) scale (34), hence the regression results for the 15% cut-off point are presented (to note: the regression results for the other cut-off points can be found in the supplementary files (supplementary file – Table 6-19)). At the 15% cut-off, prevalence of cases ≤4 months was 5.9, 4.3, 6.9 and 7.9 percentage point higher compared to those uninfected (prevalence cases ≤4 months minus prevalence uninfected) for SF-6D, MCS, PCS and CIS respectively (supplementary file – Table 2-5). In the multivariable logistic regression, significant differences between those uninfected and cases ≤4 months were observed for health utility (OR [CI]: 1.60 [1.15-2.18]), physical health (OR [CI]: 1.92 [1.45-2.53]) and fatigue (OR [CI]: 1.63 [1.25-2.12]), but not for mental health (OR [CI]: 1.19 [0.88-1.57]) (Table 2). For all outcomes, there was no significant difference observed between those uninfected and cases >4 months at the 15% cut-off point (or any of the other cut-off points).

**Table 2.**
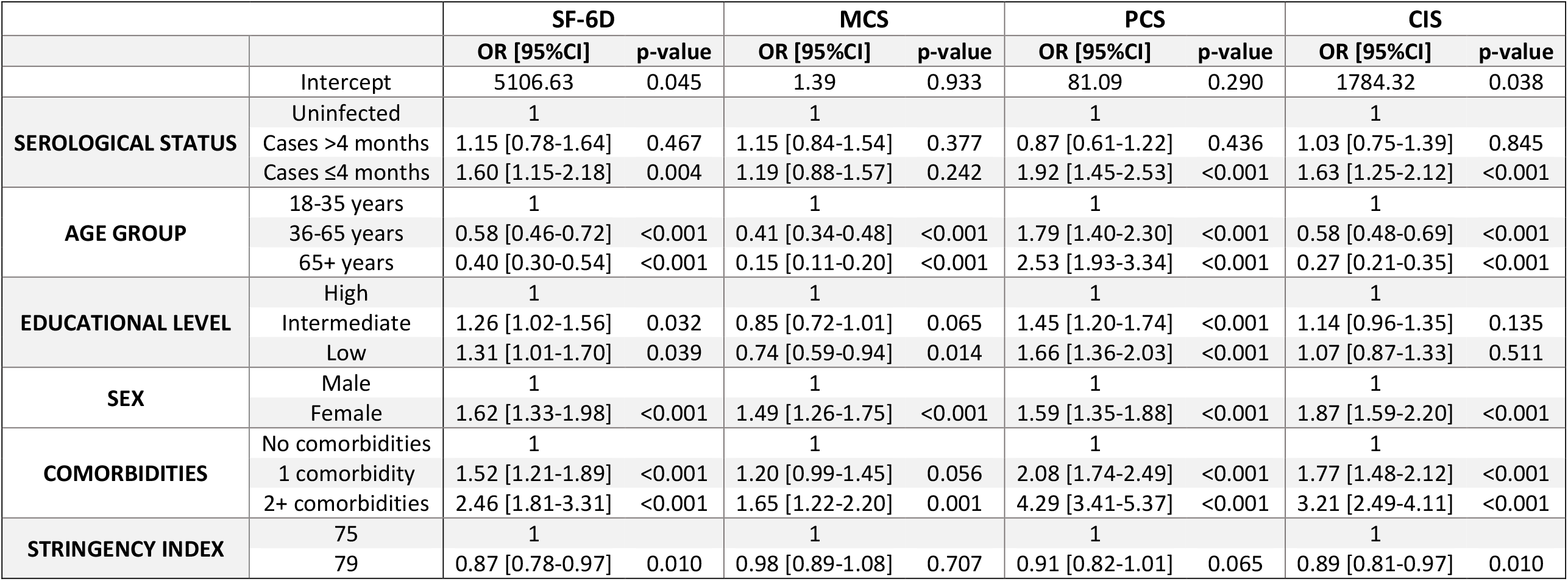
Multivariable logistic regression results for SF-6D, MCS, PCS and CIS at the 15% cut-off.

At the 15% cut-off point, multivariable logistic regressions (Table 2) showed that age groups 35-65 and 66+ years were significantly associated with poorer physical health, but with better health utility, mental health, and fatigue (p<0.001) compared to age group 18-35 years. Female sex (p<0.001) and having comorbidities (p<0.001 to p=0.001) were significantly associated with poorer outcomes on all four scales, except for mental health and having one comorbidity (p=0.001). Low educational level was significantly associated with better mental health (p=0.014), but poorer health utility (p=0.039) and physical health (p<0.001). A higher stringency index was significantly associated with better health utility and fatigue (p=0.010).

### Post-hoc analysis

In the post-hoc analysis we investigated the power to detect a minimal difference in prevalence between cases >4 months and those uninfected required to detect a significant difference (with those uninfected as reference group). Given the 327 cases >4 months, a difference in prevalence at the 15% cut-off would only become significant when the prevalence in this group was at least 4.1% points higher than that of the uninfected for the SF-6D, 5.5% for the MCS, 3.3% for the PCS and 5.2% for the CIS (supplementary file – Figure 1-4). Using the given cut-offs, the largest possible difference while having a non-significant outcome of the logistic regression would be 7.3%, 7.4%, 4.7% or 7.0% respectively.

## Discussion

In this nationwide Dutch cohort study, we assessed the relationship between time since serologically identified SARS-CoV-2 infection and HRQoL (health utility, physical health, mental health) and fatigue among unvaccinated individuals in the first year of the COVID-19 pandemic. Our findings show that recently infected participants (≤4 months ago) more often had poor outcomes in health utility, physical health and fatigue compared to those uninfected (at the 15% cut-off). Almost no significant differences were observed for mental health. For participants infected more than 4 months ago, no statistically significant differences were found compared to those uninfected, though post-hoc analyses showed a power to detect prevalence differences as low as 7%.

These observations are interesting, as they suggest that at the population level (including those symptomatic and asymptomatic) the impact of long-term sequelae after a SARS-CoV-2 infection was lower in our sample compared to the lowest estimated PCC prevalence as currently found in literature (9%) (10). The limited impact that we measured may be explained by a combination of factors. Firstly, in this study we focused on predominantly wild-type SARS-CoV-2 infected individuals, since the Alpha variant of concern started circulating in the end of 2020 and was not dominant until week 7 of 2021 (15-21 February 2021). With the onset of the Alpha variant coinciding with the beginning of the Dutch vaccination campaign for susceptible populations, disentangling the effects of the variant, age and vaccination on PCC endpoints becomes difficult. As a result, there is no evident consensus on the between-variant differences for PCC prior to Omicron as well as the influence of vaccination on PCC development and progression. The definition used, population studied, and variation in vaccination status resulted in large heterogeneity (14). Some studies report that the wild-type SARS-CoV-2 variant is associated with similar odds of long-term symptoms as the Alpha or Delta variants, after adjusting for vaccination status (37, 38). On the other hand, other vaccination-controlled evidence suggests that early pandemic variants held higher risk of PCC (14, 39). A large U.S. cohort study revealed a significant decrease in PCC incidence from pre-Delta to Omicron, primarily attributable to vaccination, with minor contributions from time-related factors such as variants (40). Secondly, our approach of serological SARS-CoV-2 detection is superior to methodologies used in many other studies (e.g. diagnostic SARS-CoV-2 testing) in terms of sensitivity and specificity, given the high test specifics of our assay (23, 41). This enabled us to include mild and asymptomatically infected individuals, who had milder acute COVID-19 and subsequently may have had lower risk for PCC (42). Thirdly, the individuals at risk for severe COVID-19, such as those with underlying conditions, may have also been at risk for long-term health impacts. However, they were less likely to be infected due to adapted behaviors (26, 43, 44). Since our study was randomly sampled from the Dutch population, the impact of several risk factors such as hospitalization, admission to intensive care unit, having comorbidities, smoking and high body mass index (45, 46) are less visible from our sample. This is because the majority of our participants experienced mild infections, and risk groups were not overrepresented in our population-based sample. Fourthly, because of the sampling, there is variability in time since infection for those infected more than 4 months ago. The majority of these cases are likely to have occurred in March and April 2020, given the epidemic curve in the Netherlands before September 2020. This equates to 10 to 11 months since infection, which is long and can contribute to the lower observed prevalence. Further subdivision of our cases >4 months into exact round of seroconversion was not possible due to the small numbers (PICO1: N=64; PICO2: N=40; PICO3: N=20).

We observed differences in health utility between those uninfected and cases ≤4 months, which is in line with the study by Poudel (2021). Poudel (2021) reported that quality of life was impacted more during the acute phase of SARS-CoV-2 infection (47). Although HRQoL can also be impacted among PCC patients, we did not observe any significant differences in health utility between those uninfected and cases >4 months. There were no differences in mental health observed in our study between those uninfected and cases >4 months or ≤4 months. This could be because the severe restrictions that were in place in February 2021 (schools were closed, people had to work from home and an evening curfew was in place from January 20^th^ until April 28^th^) had an impact on the entire population, irrespective of infection status. Literature also shows that although society as a whole was affected by the pandemic, some groups in society were disproportionally affected such as females and younger age groups (48). On the other hand, in a study examining HRQoL over the pandemic years, we did not observe a relationship between quality of life and the stringency of non-pharmaceutical interventions, nor did we find an indication that HRQoL was very different in February 2021 compared to the period after (49).

There are some limitations to consider in the interpretation of our results. Firstly, because we use serological data, acute infections with SARS-CoV-2 just before blood sampling might not have been detected in our analysis as it can take two to three weeks to seroconvert after primary infection (50). Similarly, some individuals may have been misclassified in the cases or uninfected groups due to false negative or false positive serological testing. Few individuals do not seroconvert after infection (51–56). To minimize the effect of misclassification on our estimation, we excluded participants that reported a positive PCR or rapid antigen test from the uninfected group. However, since widescale PCR testing was not available in the Netherlands until June 2020, there is still a possibility of misclassification. Given that the validation studies of the assay used showed a high sensitivity and specificity (23, 41), we believe these consist of a small minority, and hence the overall impact on our results is limited. Moreover, the use of serological data to identify cases enabled identification of infections, independent of testing accessibility, policies and adherence, which is a major strength of this study. Secondly, there might be some heterogeneity in SARS-CoV-2 infection variants in our cases ≤4 months. Given that the Alpha variant of concern started circulating in the end of 2020, but was not dominant until week 7 of 2021 (15-21 February 2021), we expect this to only be a minority of infections and thus only marginally impact our results (57). Thirdly, reinfections were not taken into account in our study. However, as very few reinfections occurred early in the pandemic (58), this most likely did not affect our results (59, 60). Finally, in this study we included a study population that was randomly selected from the Dutch population in order to ascertain representativeness of the Dutch population. However, despite these efforts, participants of Dutch descent and potentially more health-oriented persons were overrepresented, a phenomenon commonly seen in other studies as well. In addition, since the vaccination rollout in the Netherlands initially prioritized vulnerable groups and healthcare workers, the exclusion of vaccinated individuals in our study may have affected the representativeness of our results.

Focusing on early-pandemic serologically identified infections among unvaccinated individuals provides valuable insights into the overall temporal health dynamics following SARS-CoV-2 infection in a largely naïve population. With waning vaccine-induced immunity and discontinued testing policy, these results may reflect current PCC susceptibility among symptomatic and asymptomatic SARS-CoV-2 infections. In addition, they may serve as a benchmark for PCC estimates following infections with later SARS-CoV-2 variants in more immunologically heterogeneous populations. Our results could also shed light into suspectibility to other respiratory infections for which the population is largely naïve (37, 61, 62).

## Conclusion

In a Dutch randomly-selected population-based seroepidemiological cohort of SARS-CoV-2 infected unvaccinated individuals, those infected in the previous four months more often reported poor health utility and physical health, and were more often severely fatigued compared to those uninfected. HRQoL and fatigue remained below the detection limit for those infected more than four months ago, suggesting a relatively low prevalence of PCC.

## Acknowledgements

We would like to thank our colleagues from the National Institute of Public Health and Environment (RIVM), Centre of Infectious Disease Control for their contributions regarding logistics, laboratory analyses, methodological insights and manuscript reviewing.

This work was supported by the Ministry of Health, Welfare and Sports (VWS), the Netherlands. The funders had no role in study design, data collection and analysis, decision to publish, or preparation of the manuscript. There was no additional external funding received for this study.

## Data availability

The data that support the findings of this study can be requested via the PICO study website (https://www.rivm.nl/en/pienter-corona-study/information-for-researchers). Restrictions may apply to the availability of these data.

