## Supplementary file for "The impact of SARS-CoV-2 on fatigue and quality of life in the early phase of the pandemic: prevalence of post COVID-19 condition among unvaccinated individuals in a Dutch population-based serosurveillance cohort"

Table 1. Demographic and health characteristics of all included participants*

|  | UNINFECTED | Cases >4 months | Cases ≤4 months |
| --- | --- | --- | --- |
| *N* | 4,674 | 337 | 364 |
| Sex = Female *% (n)* | 55.4 (2,588) | 56.1 (189) | 58.2 (212) |
| Age groups *% (n)* |  |  |  |
| 15-35 | 21.5 (1,005) | 28.8 (97) | 32.4 (118) |
| 36-65 | 51.7 (2,417) | 47.8 (161) | 50.5 (184) |
| 66+ | 26.8 (1,252) | 23.4 (79) | 17.0 (62) |
| Educational level^A^ *% (n)* |  |  |  |
| High | 46.7 (2,184) | 42.4 (143) | 37.4 (136) |
| Intermediate | 31.2 (1,459) | 36.8 (124) | 38.7 (141) |
| Low | 20.1 (939) | 17.8 (60) | 20.9 (76) |
| Missing | 2.0 (92) | 3.0 (10) | 3.0 (11) |
| Number of comorbidities^B^ *% (n)* |  |  |  |
| None | 64.9 (3,035) | 63.5 (214) | 71.4 (260) |
| One | 24.3 (1,136) | 26.4 (89) | 19.2 (70) |
| Two or more | 9.3 (434) | 7.7 (26) | 7.1 (26) |
| MIssing | 1.5 (69) | 2.4 (8) | 2.2 (8) |
| Health utility (SF-6D) *mean (SD)* | 0.82 (0.11) | 0.81 (0.11) | 0.79 (0.13) |
| Physical health (PCS) *mean (SD)* | 54.51 (7.56) | 54.94 (6.80) | 53.13 (8.73) |
| Mental health (MCS) *mean (SD)* | 47.84 (10.27) | 47.26 (11.04) | 46.28 (10.87) |
| Fatigue (CIS) *mean (SD)* | 21.70 (11.67) | 21.72 (11.36) | 24.79 (12.89) |

* All characteristics were determined in PICO4, except for educational level and comorbidities (determined at baseline).

^A^ Educational level was classified as low (no education or primary education), intermediate (secondary school or vocational training), or high (bachelor’s degree, university).

^B^ Included comorbidities are asthma, cancer, cardiovascular disease, diabetes (type 1 & 2), immunodeficiency, pulmonary disease and renal disease.

Table 2. Proportion of health utility scores (SF-6D) in all three study groups at all cut-off points and corresponding SF-6D score.

| Cut-off point | Corresponding SF-6D score | Uninfected | Cases >4 months | Cases ≤4 months |
| --- | --- | --- | --- | --- |
| 5% | 0.603 | 0.056 | 0.067 | 0.091 |
| 10% | 0.657 | 0.126 | 0.138 | 0.194 |
| 15% | 0.660 | 0.158 | 0.171 | 0.217 |
| 20% | 0.723 | 0.221 | 0.235 | 0.282 |
| 25% | 0.782 | 0.251 | 0.263 | 0.325 |
| 30% | 0.797 | 0.305 | 0.315 | 0.379 |
| 35% | 0.800 | 0.441 | 0.468 | 0.516 |
| 40% | 0.800 | 0.441 | 0.468 | 0.516 |
| 45% | 0.856 | 0.452 | 0.480 | 0.516 |
| 50% | 0.863 | 0.760 | 0.743 | 0.769 |
| 55% | 0.863 | 0.760 | 0.743 | 0.769 |
| 60% | 0.863 | 0.760 | 0.743 | 0.769 |
| 65% | 0.863 | 0.760 | 0.743 | 0.769 |
| 70% | 0.863 | 0.760 | 0.743 | 0.769 |
| 75% | 0.863 | 0.760 | 0.743 | 0.769 |

Table 3. Proportion of mental health scores (MCS) in all three study groups at all cut-off points and corresponding MCS score.

| Cut-off point | Corresponding MCS score | Uninfected | Cases >4 months | Cases ≤4 months |
| --- | --- | --- | --- | --- |
| 5% | 25.250 | 0.050 | 0.070 | 0.063 |
| 10% | 31.828 | 0.100 | 0.116 | 0.128 |
| 15% | 36.766 | 0.151 | 0.180 | 0.194 |
| 20% | 41.105 | 0.200 | 0.226 | 0.248 |
| 25% | 43.743 | 0.250 | 0.266 | 0.305 |
| 30% | 45.892 | 0.300 | 0.315 | 0.379 |
| 35% | 47.353 | 0.351 | 0.355 | 0.416 |
| 40% | 48.721 | 0.403 | 0.404 | 0.476 |
| 45% | 49.951 | 0.451 | 0.453 | 0.527 |
| 50% | 50.675 | 0.506 | 0.514 | 0.570 |
| 55% | 51.497 | 0.550 | 0.541 | 0.601 |
| 60% | 52.513 | 0.602 | 0.609 | 0.658 |
| 65% | 53.077 | 0.651 | 0.651 | 0.687 |
| 70% | 53.727 | 0.700 | 0.682 | 0.729 |
| 75% | 55.145 | 0.758 | 0.780 | 0.792 |

Table 4. Proportion of physical health scores (PCS) in all three study groups at all cut-off points and corresponding PCS score.

| Cut-off point | Corresponding PCS score | Uninfected | Cases >4 months | Cases ≤4 months |
| --- | --- | --- | --- | --- |
| 5% | 38.004 | 0.050 | 0.037 | 0.080 |
| 10% | 44.404 | 0.100 | 0.086 | 0.160 |
| 15% | 48.359 | 0.150 | 0.128 | 0.219 |
| 20% | 51.215 | 0.201 | 0.187 | 0.293 |
| 25% | 52.515 | 0.250 | 0.248 | 0.339 |
| 30% | 54.178 | 0.300 | 0.300 | 0.385 |
| 35% | 55.226 | 0.350 | 0.339 | 0.427 |
| 40% | 55.703 | 0.403 | 0.410 | 0.467 |
| 45% | 56.160 | 0.450 | 0.443 | 0.496 |
| 50% | 56.794 | 0.514 | 0.498 | 0.541 |
| 55% | 57.262 | 0.577 | 0.566 | 0.590 |
| 60% | 57.506 | 0.602 | 0.581 | 0.613 |
| 65% | 58.014 | 0.651 | 0.642 | 0.647 |
| 70% | 58.222 | 0.709 | 0.688 | 0.684 |
| 75% | 58.752 | 0.768 | 0.755 | 0.741 |

Table 5. Proportion of fatigue scores (CIS) in all three study groups at all cut-off points and corresponding CIS score. Since higher CIS scores indicate worse outcomes, proportions have been inverted similarly to figure 5.

| Cut-off point | Corresponding CIS score | Uninfected | Cases >4 months | Cases ≤4 months |
| --- | --- | --- | --- | --- |
| 75% | 12.000 | 0.724 | 0.731 | 0.781 |
| 70% | 13.000 | 0.694 | 0.694 | 0.755 |
| 65% | 15.000 | 0.618 | 0.627 | 0.681 |
| 60% | 16.000 | 0.582 | 0.590 | 0.658 |
| 55% | 18.000 | 0.521 | 0.529 | 0.618 |
| 50% | 19.000 | 0.492 | 0.514 | 0.595 |
| 45% | 21.000 | 0.436 | 0.453 | 0.519 |
| 40% | 23.000 | 0.383 | 0.407 | 0.487 |
| 35% | 25.000 | 0.332 | 0.343 | 0.459 |
| 30% | 27.000 | 0.287 | 0.281 | 0.402 |
| 25% | 29.000 | 0.248 | 0.257 | 0.362 |
| 20% | 32.000 | 0.186 | 0.187 | 0.291 |
| 15% | 35.000 | 0.146 | 0.150 | 0.225 |
| 10% | 39.000 | 0.094 | 0.092 | 0.145 |
| 5% | 44.000 | 0.049 | 0.043 | 0.088 |

Table 6. Results of the four multivariable logistic regression models for health utility (SF-6D), mental health (MCS), physical health (PCS) and fatigue (CIS) at the 5% cut-off of the cumulative distribution of uninfected.

|  |  | SF-6D | | MCS | | PCS | | CIS | |
| --- | --- | --- | --- | --- | --- | --- | --- | --- | --- |
|  |  | **OR [CI]** | **p-value** | **OR [CI]** | **p-value** | **OR [CI]** | **p-value** | **OR [CI]** | **p-value** |
|  | Intercept | 5269.61 | 0.138 | 8190.66 | 0.082 | 1086.16 | 0.268 | 163193.22 | 0.014 |
| Serological status | Uninfected | 1 |  | 1 |  | 1 |  | 1 |  |
|  | Cases >4 months | 0.80 [0.42-1.40] | 0.469 | 1.29 [0.80-1.99] | 0.279 | 0.79 [0.41-1.38] | 0.434 | 1.04 [0.63-1.64] | 0.865 |
|  | Cases ≤4 months | 1.44 [0.89-2.22] | 0.116 | 1.06 [0.65-1.65] | 0.797 | 2.11 [1.34-3.19] | 0.001 | 1.76 [1.18-2.53] | 0.004 |
| Age group | 18-35 years | 1 |  | 1 |  | 1 |  | 1 |  |
|  | 36-65 years | 0.66 [0.48-0.90] | 0.009 | 0.35 [0.27-0.46] | <0.001 | 3.22 [1.93-5.79] | <0.001 | 0.65 [0.49-0.85] | 0.002 |
|  | 65+ years | 0.37 [0.23-0.56] | <0.001 | 0.10 [0.06-0.18] | <0.001 | 3.67 [2.12-6.77] | <0.001 | 0.34 [0.23-0.50] | <0.001 |
| Educational level | High | 1 |  | 1 |  | 1 |  | 1 |  |
|  | Intermediate | 1.18 [0.86-1.60] | 0.303 | 0.92 [0.70-1.20] | 0.529 | 1.31 [0.95-1.80] | 0.099 | 1.22 [0.94-1.59] | 0.137 |
|  | Low | 1.61 [1.13-2.27] | 0.008 | 0.78 [0.52-1.15] | 0.232 | 2.07 [1.51-2.85] | <0.001 | 1.32 [0.96-1.82] | 0.085 |
| Sex | Male | 1 |  | 1 |  | 1 |  | 1 |  |
|  | Female | 1.60 [1.21-2.13] | 0.001 | 1.99 [1.52-2.64] | <0.001 | 1.52 [1.17-1.99] | 0.002 | 2.11 [1.64-2.75] | <0.001 |
| Comorbidities | No comorbidities | 1 |  | 1 |  | 1 |  | 1 |  |
|  | 1 comorbidity | 1.59 [1.15-2.17] | 0.004 | 1.31 [0.96-1.76] | 0.084 | 2.52 [1.85-3.42] | <0.001 | 1.75 [1.33-2.30] | <0.001 |
|  | 2+ comorbidities | 3.28 [2.20-4.81] | <0.001 | 1.65 [0.97-2.65] | 0.050 | 6.51 [4.68-9.06] | <0.001 | 3.43 [2.39-4.86] | <0.001 |
| Stringency index | 75 | 1 |  | 1 |  | 1 |  | 1 |  |
|  | 79 | 0.86 [0.75-1.00] | 0.039 | 0.86 [0.76-0.99] | 0.026 | 0.86 [0.74-1.01] | 0.054 | 0.82 [0.73-0.94] | 0.002 |

Table 7. Results of the four multivariable logistic regression models for health utility (SF-6D), mental health (MCS), physical health (PCS) and fatigue (CIS) at the 10% cut-off of the cumulative distribution of uninfected.

|  |  | SF-6D | | MCS | | PCS | | CIS | |
| --- | --- | --- | --- | --- | --- | --- | --- | --- | --- |
|  |  | **OR [CI]** | **p-value** | **OR [CI]** | **p-value** | **OR [CI]** | **p-value** | **OR [CI]** | **p-value** |
|  | Intercept | 5372.20 | 0.044 | 21.61 | 0.478 | 894.67 | 0.147 | 36445.55 | 0.008 |
| Serological status | Uninfected | 1 |  | 1 |  | 1 |  | 1 |  |
|  | Cases >4 months | 1.15 [0.78-1.64] | 0.467 | 1.07 [0.73-1.51] | 0.721 | 0.90 [0.58-1.33] | 0.607 | 0.85 [0.57-1.23] | 0.413 |
|  | Cases ≤4 months | 1.56 [1.12-2.13] | 0.007 | 1.13 [0.80-1.57] | 0.479 | 2.10 [1.52-2.87] | <0.001 | 1.39 [1.00-1.89] | 0.042 |
| Age group | 18-35 years | 1 |  | 1 |  | 1 |  | 1 |  |
|  | 36-65 years | 0.57 [0.46-0.72] | <0.001 | 0.41 [0.33-0.49] | <0.001 | 2.12 [1.56-2.93] | <0.001 | 0.64 [0.52-0.79] | <0.001 |
|  | 65+ years | 0.40 [0.30-0.54] | <0.001 | 0.13 [0.09-0.19] | <0.001 | 2.77 [1.97-3.93] | <0.001 | 0.27 [0.20-0.37] | <0.001 |
| Educational level | High | 1 |  | 1 |  | 1 |  | 1 |  |
|  | Intermediate | 1.25 [1.01-1.55] | 0.037 | 0.99 [0.81-1.21] | 0.916 | 1.48 [1.19-1.85] | <0.001 | 1.16 [0.95-1.42] | 0.136 |
|  | Low | 1.31 [1.01-1.70] | 0.039 | 0.77 [0.57-1.02] | 0.074 | 1.76 [1.39-2.23] | <0.001 | 1.16 [0.90-1.49] | 0.235 |
| Sex | Male | 1 |  | 1 |  | 1 |  | 1 |  |
|  | Female | 1.63 [1.34-2.00] | <0.001 | 1.59 [1.31-1.93] | <0.001 | 1.78 [1.47-2.17] | <0.001 | 1.85 [1.53-2.25] | <0.001 |
| Comorbidities | No comorbidities | 1 |  | 1 |  | 1 |  | 1 |  |
|  | 1 comorbidity | 1.52 [1.22-1.90] | <0.001 | 1.23 [0.98-1.54] | 0.066 | 2.18 [1.75-2.69] | <0.001 | 1.75 [1.42-2.16] | <0.001 |
|  | 2+ comorbidities | 2.47 [1.82-3.33] | <0.001 | 1.68 [1.17-2.37] | 0.004 | 4.55 [3.52-5.87] | <0.001 | 3.17 [2.38-4.21] | <0.001 |
| Stringency index | 75 | 1 |  | 1 |  | 1 |  | 1 |  |
|  | 79 | 0.87 [0.78-0.97] | 0.010 | 0.94 [0.85-1.05] | 0.270 | 0.87 [0.78-0.99] | 0.022 | 0.85 [0.77-0.94] | 0.001 |

Table 8. Results of the four multivariable logistic regression models for health utility (SF-6D), mental health (MCS), physical health (PCS) and fatigue (CIS) at the 20% cut-off of the cumulative distribution of uninfected.

|  |  | SF-6D | | MCS | | PCS | | CIS | |
| --- | --- | --- | --- | --- | --- | --- | --- | --- | --- |
|  |  | **OR [CI]** | **p-value** | **OR [CI]** | **p-value** | **OR [CI]** | **p-value** | **OR [CI]** | **p-value** |
|  | Intercept | 141.29 | 0.155 | 0.65 | 0.905 | 20.93 | 0.425 | 91.68 | 0.186 |
| Serological status | Uninfected | 1 |  | 1 |  | 1 |  | 1 |  |
|  | Cases >4 months | 1.00 [0.75-1.33] | 0.973 | 1.08 [0.81-1.42] | 0.604 | 0.96 [0.71-1.29] | 0.806 | 1.02 [0.77-1.35] | 0.880 |
|  | Cases ≤4 months | 1.32 [1.01-1.70] | 0.039 | 1.16 [0.89-1.51] | 0.263 | 2.02 [1.56-2.59] | <0.001 | 1.67 [1.31-2.14] | <0.001 |
| Age group | 18-35 years | 1 |  | 1 |  | 1 |  | 1 |  |
|  | 36-65 years | 0.49 [0.41-0.58] | <0.001 | 0.41 [0.35-0.48] | <0.001 | 1.61 [1.30-1.99] | <0.001 | 0.57 [0.48-0.67] | <0.001 |
|  | 65+ years | 0.31 [0.25-0.39] | <0.001 | 0.16 [0.12-0.20] | <0.001 | 2.48 [1.96-3.14] | <0.001 | 0.27 [0.22-0.34] | <0.001 |
| Educational level | High | 1 |  | 1 |  | 1 |  | 1 |  |
|  | Intermediate | 0.92 [0.79-1.08] | 0.319 | 0.79 [0.67-0.92] | 0.003 | 1.44 [1.22-1.69] | <0.001 | 1.07 [0.91-1.24] | 0.411 |
|  | Low | 0.89 [0.73-1.09] | 0.253 | 0.69 [0.56-0.85] | 0.001 | 1.63 [1.35-1.95] | <0.001 | 0.96 [0.79-1.17] | 0.705 |
| Sex | Male | 1 |  | 1 |  | 1 |  | 1 |  |
|  | Female | 1.47 [1.27-1.70] | <0.001 | 1.41 [1.22-1.63] | <0.001 | 1.57 [1.36-1.82] | <0.001 | 1.63 [1.41-1.88] | <0.001 |
| Comorbidities | No comorbidities | 1 |  | 1 |  | 1 |  | 1 |  |
|  | 1 comorbidity | 1.59 [1.34-1.88] | <0.001 | 1.23 [1.03-1.46] | 0.018 | 2.00 [1.70-2.34] | <0.001 | 1.67 [1.41-1.96] | <0.001 |
|  | 2+ comorbidities | 2.57 [2.02-3.25] | <0.001 | 1.57 [1.20-2.04] | 0.001 | 4.10 [3.31-5.08] | <0.001 | 3.44 [2.73-4.32] | <0.001 |
| Stringency index | 75 | 1 |  | 1 |  | 1 |  | 1 |  |
|  | 79 | 0.92 [0.85-1.01] | 0.076 | 1.00 [0.91-1.09] | 0.938 | 0.93 [0.85-1.02] | 0.126 | 0.93 [0.85-1.01] | 0.086 |

Table 9. Results of the four multivariable logistic regression models for health utility (SF-6D), mental health (MCS), physical health (PCS) and fatigue (CIS) at the 25% cut-off of the cumulative distribution of uninfected.

|  |  | SF-6D | | MCS | | PCS | | CIS | |
| --- | --- | --- | --- | --- | --- | --- | --- | --- | --- |
|  |  | **OR [CI]** | **p-value** | **OR [CI]** | **p-value** | **OR [CI]** | **p-value** | **OR [CI]** | **p-value** |
|  | Intercept | 68.83 | 0.194 | 2.70 | 0.766 | 6.42 | 0.603 | 324.23 | 0.069 |
| Serological status | Uninfected | 1 |  | 1 |  | 1 |  | 1 |  |
|  | Cases >4 months | 1.01 [0.78-1.31] | 0.926 | 0.99 [0.76-1.29] | 0.969 | 1.03 [0.78-1.35] | 0.821 | 0.91 [0.70-1.18] | 0.505 |
|  | Cases ≤4 months | 1.36 [1.07-1.73] | 0.012 | 1.17 [0.91-1.49] | 0.228 | 1.83 [1.43-2.34] | <0.001 | 1.55 [1.22-1.96] | <0.001 |
| Age group | 18-35 years | 1 |  | 1 |  | 1 |  | 1 |  |
|  | 36-65 years | 0.46 [0.39-0.53] | <0.001 | 0.39 [0.34-0.46] | <0.001 | 1.67 [1.38-2.02] | <0.001 | 0.58 [0.50-0.68] | <0.001 |
|  | 65+ years | 0.29 [0.23-0.35] | <0.001 | 0.17 [0.14-0.22] | <0.001 | 2.40 [1.94-2.99] | <0.001 | 0.29 [0.24-0.35] | <0.001 |
| Educational level | High | 1 |  | 1 |  | 1 |  | 1 |  |
|  | Intermediate | 0.89 [0.77-1.03] | 0.108 | 0.79 [0.69-0.92] | 0.002 | 1.47 [1.27-1.71] | <0.001 | 1.00 [0.87-1.15] | 0.971 |
|  | Low | 0.90 [0.75-1.08] | 0.251 | 0.68 [0.56-0.83] | <0.001 | 1.71 [1.44-2.02] | <0.001 | 0.97 [0.82-1.16] | 0.778 |
| Sex | Male | 1 |  | 1 |  | 1 |  | 1 |  |
|  | Female | 1.43 [1.26-1.64] | <0.001 | 1.47 [1.29-1.68] | <0.001 | 1.53 [1.34-1.76] | <0.001 | 1.65 [1.45-1.88] | <0.001 |
| Comorbidities | No comorbidities | 1 |  | 1 |  | 1 |  | 1 |  |
|  | 1 comorbidity | 1.51 [1.29-1.76] | <0.001 | 1.27 [1.08-1.48] | 0.003 | 2.08 [1.80-2.42] | <0.001 | 1.70 [1.46-1.97] | <0.001 |
|  | 2+ comorbidities | 2.50 [2.00-3.12] | <0.001 | 1.38 [1.07-1.77] | 0.011 | 3.97 [3.22-4.89] | <0.001 | 3.18 [2.56-3.96] | <0.001 |
| Stringency index | 75 | 1 |  | 1 |  | 1 |  | 1 |  |
|  | 79 | 0.94 [0.87-1.02] | 0.126 | 0.98 [0.91-1.07] | 0.673 | 0.95 [0.87-1.04] | 0.224 | 0.92 [0.85-0.99] | 0.034 |

Table 10. Results of the four multivariable logistic regression models for health utility (SF-6D), mental health (MCS), physical health (PCS) and fatigue (CIS) at the 30% cut-off of the cumulative distribution of uninfected.

|  |  | SF-6D | | MCS | | PCS | | CIS | |
| --- | --- | --- | --- | --- | --- | --- | --- | --- | --- |
|  |  | **OR [CI]** | **p-value** | **OR [CI]** | **p-value** | **OR [CI]** | **p-value** | **OR [CI]** | **p-value** |
|  | Intercept | 88.11 | 0.166 | 0.80 | 0.944 | 2.55 | 0.782 | 74.14 | 0.168 |
| Serological status | Uninfected | 1 |  | 1 |  | 1 |  | 1 |  |
|  | Cases >4 months | 1.01 [0.78-1.31] | 0.934 | 0.98 [0.76-1.26] | 0.892 | 1.04 [0.80-1.34] | 0.773 | 0.97 [0.75-1.24] | 0.813 |
|  | Cases ≤4 months | 1.37 [1.08-1.74] | 0.009 | 1.28 [1.01-1.62] | 0.043 | 1.70 [1.34-2.15] | <0.001 | 1.60 [1.27-2.01] | <0.001 |
| Age group | 18-35 years | 1 |  | 1 |  | 1 |  | 1 |  |
|  | 36-65 years | 0.45 [0.39-0.53] | <0.001 | 0.38 [0.33-0.44] | <0.001 | 1.61 [1.36-1.92] | <0.001 | 0.58 [0.50-0.68] | <0.001 |
|  | 65+ years | 0.29 [0.24-0.35] | <0.001 | 0.19 [0.15-0.23] | <0.001 | 2.30 [1.88-2.81] | <0.001 | 0.30 [0.25-0.37] | <0.001 |
| Educational level | High | 1 |  | 1 |  | 1 |  | 1 |  |
|  | Intermediate | 0.88 [0.77-1.02] | 0.093 | 0.79 [0.69-0.91] | 0.001 | 1.47 [1.27-1.69] | <0.001 | 0.98 [0.85-1.12] | 0.746 |
|  | Low | 0.90 [0.75-1.07] | 0.233 | 0.69 [0.58-0.82] | <0.001 | 1.56 [1.32-1.84] | <0.001 | 1.00 [0.85-1.19] | 0.967 |
| Sex | Male | 1 |  | 1 |  | 1 |  | 1 |  |
|  | Female | 1.44 [1.26-1.64] | <0.001 | 1.49 [1.31-1.69] | <0.001 | 1.57 [1.38-1.78] | <0.001 | 1.61 [1.43-1.83] | <0.001 |
| Comorbidities | No comorbidities | 1 |  | 1 |  | 1 |  | 1 |  |
|  | 1 comorbidity | 1.52 [1.31-1.77] | <0.001 | 1.25 [1.08-1.46] | 0.003 | 1.92 [1.66-2.21] | <0.001 | 1.64 [1.42-1.89] | <0.001 |
|  | 2+ comorbidities | 2.46 [1.97-3.07] | <0.001 | 1.43 [1.13-1.80] | 0.002 | 3.79 [3.08-4.66] | <0.001 | 3.13 [2.53-3.87] | <0.001 |
| Stringency index | 75 | 1 |  | 1 |  | 1 |  | 1 |  |
|  | 79 | 0.94 [0.86-1.02] | 0.108 | 1.00 [0.92-1.09] | 0.979 | 0.96 [0.88-1.05] | 0.365 | 0.94 [0.87-1.01] | 0.105 |

Table 11. Results of the four multivariable logistic regression models for health utility (SF-6D), mental health (MCS), physical health (PCS) and fatigue (CIS) at the 35% cut-off of the cumulative distribution of uninfected.

|  |  | SF-6D | | MCS | | PCS | | CIS | |
| --- | --- | --- | --- | --- | --- | --- | --- | --- | --- |
|  |  | **OR [CI]** | **p-value** | **OR [CI]** | **p-value** | **OR [CI]** | **p-value** | **OR [CI]** | **p-value** |
|  | Intercept | 88.11 | 0.166 | 2.16 | 0.806 | 0.50 | 0.833 | 231.98 | 0.075 |
| Serological status | Uninfected | 1 |  | 1 |  | 1 |  | 1 |  |
|  | Cases >4 months | 1.01 [0.78-1.31] | 0.934 | 0.93 [0.73-1.19] | 0.574 | 0.99 [0.77-1.26] | 0.907 | 1.05 [0.82-1.33] | 0.711 |
|  | Cases ≤4 months | 1.37 [1.08-1.74] | 0.009 | 1.19 [0.94-1.50] | 0.139 | 1.61 [1.28-2.03] | <0.001 | 1.53 [1.22-1.92] | <0.001 |
| Age group | 18-35 years | 1 |  | 1 |  | 1 |  | 1 |  |
|  | 36-65 years | 0.45 [0.39-0.53] | <0.001 | 0.38 [0.33-0.44] | <0.001 | 1.59 [1.35-1.87] | <0.001 | 0.55 [0.48-0.64] | <0.001 |
|  | 65+ years | 0.29 [0.24-0.35] | <0.001 | 0.19 [0.16-0.23] | <0.001 | 2.50 [2.07-3.03] | <0.001 | 0.28 [0.24-0.34] | <0.001 |
| Educational level | High | 1 |  | 1 |  | 1 |  | 1 |  |
|  | Intermediate | 0.88 [0.77-1.02] | 0.093 | 0.83 [0.73-0.95] | 0.006 | 1.52 [1.33-1.74] | <0.001 | 0.99 [0.87-1.13] | 0.867 |
|  | Low | 0.90 [0.75-1.07] | 0.233 | 0.73 [0.61-0.86] | <0.001 | 1.62 [1.38-1.90] | <0.001 | 0.99 [0.84-1.16] | 0.898 |
| Sex | Male | 1 |  | 1 |  | 1 |  | 1 |  |
|  | Female | 1.44 [1.26-1.64] | <0.001 | 1.49 [1.32-1.68] | <0.001 | 1.49 [1.32-1.69] | <0.001 | 1.66 [1.47-1.87] | <0.001 |
| Comorbidities | No comorbidities | 1 |  | 1 |  | 1 |  | 1 |  |
|  | 1 comorbidity | 1.52 [1.31-1.77] | <0.001 | 1.23 [1.07-1.42] | 0.005 | 1.91 [1.67-2.20] | <0.001 | 1.72 [1.49-1.98] | <0.001 |
|  | 2+ comorbidities | 2.46 [1.97-3.07] | <0.001 | 1.32 [1.06-1.65] | 0.014 | 3.61 [2.94-4.46] | <0.001 | 3.14 [2.54-3.87] | <0.001 |
| Stringency index | 75 | 1 |  | 1 |  | 1 |  | 1 |  |
|  | 79 | 0.94 [0.86-1.02] | 0.108 | 0.99 [0.92-1.07] | 0.825 | 0.98 [0.91-1.07] | 0.718 | 0.93 [0.86-1.00] | 0.052 |

Table 12. Results of the four multivariable logistic regression models for health utility (SF-6D), mental health (MCS), physical health (PCS) and fatigue (CIS) at the 40% cut-off of the cumulative distribution of uninfected.

|  |  | SF-6D | | MCS | | PCS | | CIS | |
| --- | --- | --- | --- | --- | --- | --- | --- | --- | --- |
|  |  | **OR [CI]** | **p-value** | **OR [CI]** | **p-value** | **OR [CI]** | **p-value** | **OR [CI]** | **p-value** |
|  | Intercept | 88.11 | 0.166 | 1.46 | 0.902 | 0.41 | 0.784 | 454.34 | 0.044 |
| Serological status | Uninfected | 1 |  | 1 |  | 1 |  | 1 |  |
|  | Cases >4 months | 1.01 [0.78-1.31] | 0.934 | 0.90 [0.70-1.14] | 0.387 | 1.05 [0.82-1.33] | 0.719 | 1.02 [0.80-1.29] | 0.869 |
|  | Cases ≤4 months | 1.37 [1.08-1.74] | 0.009 | 1.19 [0.95-1.50] | 0.135 | 1.53 [1.21-1.92] | <0.001 | 1.34 [1.06-1.67] | 0.012 |
| Age group | 18-35 years | 1 |  | 1 |  | 1 |  | 1 |  |
|  | 36-65 years | 0.45 [0.39-0.53] | <0.001 | 0.37 [0.32-0.42] | <0.001 | 1.64 [1.40-1.92] | <0.001 | 0.52 [0.45-0.60] | <0.001 |
|  | 65+ years | 0.29 [0.24-0.35] | <0.001 | 0.19 [0.15-0.22] | <0.001 | 2.72 [2.27-3.28] | <0.001 | 0.27 [0.23-0.33] | <0.001 |
| Educational level | High | 1 |  | 1 |  | 1 |  | 1 |  |
|  | Intermediate | 0.88 [0.77-1.02] | 0.093 | 0.84 [0.74-0.96] | 0.009 | 1.53 [1.34-1.74] | <0.001 | 0.97 [0.85-1.11] | 0.659 |
|  | Low | 0.90 [0.75-1.07] | 0.233 | 0.77 [0.66-0.91] | 0.002 | 1.70 [1.46-1.99] | <0.001 | 0.96 [0.82-1.12] | 0.572 |
| Sex | Male | 1 |  | 1 |  | 1 |  | 1 |  |
|  | Female | 1.44 [1.26-1.64] | <0.001 | 1.48 [1.31-1.66] | <0.001 | 1.39 [1.23-1.56] | <0.001 | 1.67 [1.48-1.87] | <0.001 |
| Comorbidities | No comorbidities | 1 |  | 1 |  | 1 |  | 1 |  |
|  | 1 comorbidity | 1.52 [1.31-1.77] | <0.001 | 1.19 [1.03-1.37] | 0.015 | 1.82 [1.59-2.09] | <0.001 | 1.62 [1.41-1.86] | <0.001 |
|  | 2+ comorbidities | 2.46 [1.97-3.07] | <0.001 | 1.34 [1.08-1.66] | 0.007 | 3.59 [2.90-4.46] | <0.001 | 3.11 [2.52-3.83] | <0.001 |
| Stringency index | 75 | 1 |  | 1 |  | 1 |  | 1 |  |
|  | 79 | 0.94 [0.86-1.02] | 0.108 | 1.00 [0.93-1.08] | 0.989 | 0.99 [0.91-1.07] | 0.810 | 0.92 [0.86-1.00] | 0.038 |

Table 13. Results of the four multivariable logistic regression models for health utility (SF-6D), mental health (MCS), physical health (PCS) and fatigue (CIS) at the 45% cut-off of the cumulative distribution of uninfected.

|  |  | SF-6D | | MCS | | PCS | | CIS | |
| --- | --- | --- | --- | --- | --- | --- | --- | --- | --- |
|  |  | **OR [CI]** | **p-value** | **OR [CI]** | **p-value** | **OR [CI]** | **p-value** | **OR [CI]** | **p-value** |
|  | Intercept | 10.99 | 0.428 | 20.00 | 0.330 | 0.75 | 0.927 | 6595.21 | 0.004 |
| Serological status | Uninfected | 1 |  | 1 |  | 1 |  | 1 |  |
|  | Cases >4 months | 1.05 [0.83-1.32] | 0.702 | 0.94 [0.74-1.19] | 0.610 | 1.01 [0.80-1.28] | 0.919 | 0.97 [0.77-1.23] | 0.825 |
|  | Cases ≤4 months | 1.22 [0.98-1.53] | 0.079 | 1.20 [0.96-1.52] | 0.114 | 1.38 [1.10-1.74] | 0.005 | 1.45 [1.16-1.83] | 0.001 |
| Age group | 18-35 years | 1 |  | 1 |  | 1 |  | 1 |  |
|  | 36-65 years | 0.43 [0.37-0.50] | <0.001 | 0.37 [0.32-0.43] | <0.001 | 1.68 [1.44-1.95] | <0.001 | 0.52 [0.45-0.60] | <0.001 |
|  | 65+ years | 0.32 [0.27-0.39] | <0.001 | 0.18 [0.15-0.22] | <0.001 | 2.72 [2.27-3.26] | <0.001 | 0.26 [0.22-0.31] | <0.001 |
| Educational level | High | 1 |  | 1 |  | 1 |  | 1 |  |
|  | Intermediate | 1.01 [0.89-1.14] | 0.904 | 0.78 [0.69-0.89] | <0.001 | 1.57 [1.37-1.78] | <0.001 | 0.94 [0.82-1.07] | 0.319 |
|  | Low | 0.98 [0.84-1.15] | 0.833 | 0.76 [0.65-0.89] | 0.001 | 1.74 [1.49-2.03] | <0.001 | 0.94 [0.80-1.10] | 0.412 |
| Sex | Male | 1 |  | 1 |  | 1 |  | 1 |  |
|  | Female | 1.47 [1.31-1.65] | <0.001 | 1.52 [1.36-1.71] | <0.001 | 1.36 [1.21-1.54] | <0.001 | 1.72 [1.54-1.93] | <0.001 |
| Comorbidities | No comorbidities | 1 |  | 1 |  | 1 |  | 1 |  |
|  | 1 comorbidity | 1.46 [1.27-1.67] | <0.001 | 1.21 [1.05-1.39] | 0.008 | 1.87 [1.64-2.14] | <0.001 | 1.59 [1.38-1.82] | <0.001 |
|  | 2+ comorbidities | 1.95 [1.59-2.39] | <0.001 | 1.39 [1.13-1.71] | 0.002 | 3.85 [3.08-4.84] | <0.001 | 2.95 [2.39-3.64] | <0.001 |
| Stringency index | 75 | 1 |  | 1 |  | 1 |  | 1 |  |
|  | 79 | 0.97 [0.90-1.05] | 0.457 | 0.97 [0.90-1.05] | 0.428 | 0.99 [0.91-1.07] | 0.710 | 0.89 [0.83-0.97] | 0.005 |

Table 14. Results of the four multivariable logistic regression models for health utility (SF-6D), mental health (MCS), physical health (PCS) and fatigue (CIS) at the 50% cut-off of the cumulative distribution of uninfected.

|  |  | SF-6D | | MCS | | PCS | | CIS | |
| --- | --- | --- | --- | --- | --- | --- | --- | --- | --- |
|  |  | **OR [CI]** | **p-value** | **OR [CI]** | **p-value** | **OR [CI]** | **p-value** | **OR [CI]** | **p-value** |
|  | Intercept | 10.99 | 0.428 | 5.14 | 0.595 | 8.63 | 0.486 | 7809.64 | 0.005 |
| Serological status | Uninfected | 1 |  | 1 |  | 1 |  | 1 |  |
|  | Cases >4 months | 1.05 [0.83-1.32] | 0.702 | 0.90 [0.71-1.14] | 0.395 | 1.00 [0.79-1.27] | 0.972 | 0.97 [0.77-1.23] | 0.830 |
|  | Cases ≤4 months | 1.22 [0.98-1.53] | 0.079 | 1.16 [0.92-1.46] | 0.217 | 1.35 [1.08-1.70] | 0.010 | 1.40 [1.11-1.77] | 0.005 |
| Age group | 18-35 years | 1 |  | 1 |  | 1 |  | 1 |  |
|  | 36-65 years | 0.43 [0.37-0.50] | <0.001 | 0.36 [0.31-0.42] | <0.001 | 1.80 [1.56-2.09] | <0.001 | 0.50 [0.43-0.57] | <0.001 |
|  | 65+ years | 0.32 [0.27-0.39] | <0.001 | 0.19 [0.15-0.22] | <0.001 | 2.77 [2.32-3.33] | <0.001 | 0.25 [0.21-0.30] | <0.001 |
| Educational level | High | 1 |  | 1 |  | 1 |  | 1 |  |
|  | Intermediate | 1.01 [0.89-1.14] | 0.904 | 0.81 [0.71-0.93] | 0.002 | 1.54 [1.35-1.76] | <0.001 | 0.95 [0.83-1.08] | 0.411 |
|  | Low | 0.98 [0.84-1.15] | 0.833 | 0.76 [0.65-0.89] | 0.001 | 1.76 [1.50-2.06] | <0.001 | 0.94 [0.81-1.10] | 0.450 |
| Sex | Male | 1 |  | 1 |  | 1 |  | 1 |  |
|  | Female | 1.47 [1.31-1.65] | <0.001 | 1.55 [1.38-1.74] | <0.001 | 1.28 [1.13-1.43] | <0.001 | 1.67 [1.49-1.87] | <0.001 |
| Comorbidities | No comorbidities | 1 |  | 1 |  | 1 |  | 1 |  |
|  | 1 comorbidity | 1.46 [1.27-1.67] | <0.001 | 1.19 [1.03-1.36] | 0.016 | 1.92 [1.67-2.20] | <0.001 | 1.52 [1.33-1.75] | <0.001 |
|  | 2+ comorbidities | 1.95 [1.59-2.39] | <0.001 | 1.34 [1.09-1.65] | 0.005 | 3.96 [3.13-5.03] | <0.001 | 2.88 [2.33-3.57] | <0.001 |
| Stringency index | 75 | 1 |  | 1 |  | 1 |  | 1 |  |
|  | 79 | 0.97 [0.90-1.05] | 0.457 | 0.99 [0.92-1.07] | 0.775 | 0.96 [0.89-1.03] | 0.269 | 0.90 [0.83-0.97] | 0.007 |

Table 15. Results of the four multivariable logistic regression models for health utility (SF-6D), mental health (MCS), physical health (PCS) and fatigue (CIS) at the 55% cut-off of the cumulative distribution of uninfected.

|  |  | SF-6D | | MCS | | PCS | | CIS | |
| --- | --- | --- | --- | --- | --- | --- | --- | --- | --- |
|  |  | **OR [CI]** | **p-value** | **OR [CI]** | **p-value** | **OR [CI]** | **p-value** | **OR [CI]** | **p-value** |
|  | Intercept | 10.99 | 0.428 | 18.39 | 0.354 | 3.98 | 0.654 | 8095.92 | 0.005 |
| Serological status | Uninfected | 1 |  | 1 |  | 1 |  | 1 |  |
|  | Cases >4 months | 1.05 [0.83-1.32] | 0.702 | 0.88 [0.70-1.12] | 0.311 | 0.97 [0.77-1.24] | 0.835 | 0.94 [0.74-1.19] | 0.588 |
|  | Cases ≤4 months | 1.22 [0.98-1.53] | 0.079 | 1.08 [0.86-1.37] | 0.494 | 1.24 [0.98-1.56] | 0.072 | 1.34 [1.06-1.69] | 0.016 |
| Age group | 18-35 years | 1 |  | 1 |  | 1 |  | 1 |  |
|  | 36-65 years | 0.43 [0.37-0.50] | <0.001 | 0.36 [0.30-0.42] | <0.001 | 1.86 [1.61-2.15] | <0.001 | 0.50 [0.43-0.58] | <0.001 |
|  | 65+ years | 0.32 [0.27-0.39] | <0.001 | 0.19 [0.16-0.23] | <0.001 | 2.99 [2.50-3.59] | <0.001 | 0.24 [0.20-0.29] | <0.001 |
| Educational level | High | 1 |  | 1 |  | 1 |  | 1 |  |
|  | Intermediate | 1.01 [0.89-1.14] | 0.904 | 0.84 [0.74-0.96] | 0.010 | 1.49 [1.31-1.69] | <0.001 | 0.95 [0.83-1.08] | 0.444 |
|  | Low | 0.98 [0.84-1.15] | 0.833 | 0.78 [0.67-0.91] | 0.002 | 1.74 [1.48-2.05] | <0.001 | 0.94 [0.80-1.10] | 0.436 |
| Sex | Male | 1 |  | 1 |  | 1 |  | 1 |  |
|  | Female | 1.47 [1.31-1.65] | <0.001 | 1.56 [1.39-1.76] | <0.001 | 1.21 [1.08-1.37] | 0.001 | 1.66 [1.48-1.86] | <0.001 |
| Comorbidities | No comorbidities | 1 |  | 1 |  | 1 |  | 1 |  |
|  | 1 comorbidity | 1.46 [1.27-1.67] | <0.001 | 1.10 [0.96-1.26] | 0.182 | 1.87 [1.63-2.15] | <0.001 | 1.48 [1.29-1.70] | <0.001 |
|  | 2+ comorbidities | 1.95 [1.59-2.39] | <0.001 | 1.19 [0.97-1.46] | 0.097 | 3.72 [2.91-4.79] | <0.001 | 2.91 [2.35-3.62] | <0.001 |
| Stringency index | 75 | 1 |  | 1 |  | 1 |  | 1 |  |
|  | 79 | 0.97 [0.90-1.05] | 0.457 | 0.98 [0.90-1.06] | 0.547 | 0.97 [0.90-1.05] | 0.438 | 0.90 [0.83-0.97] | 0.008 |

Table 16. Results of the four multivariable logistic regression models for health utility (SF-6D), mental health (MCS), physical health (PCS) and fatigue (CIS) at the 60% cut-off of the cumulative distribution of uninfected.

|  |  | SF-6D | | MCS | | PCS | | CIS | |
| --- | --- | --- | --- | --- | --- | --- | --- | --- | --- |
|  |  | **OR [CI]** | **p-value** | **OR [CI]** | **p-value** | **OR [CI]** | **p-value** | **OR [CI]** | **p-value** |
|  | Intercept | 10.99 | 0.428 | 52.65 | 0.218 | 2.32 | 0.787 | 3450.16 | 0.015 |
| Serological status | Uninfected | 1 |  | 1 |  | 1 |  | 1 |  |
|  | Cases >4 months | 1.05 [0.83-1.32] | 0.702 | 0.88 [0.69-1.12] | 0.290 | 0.99 [0.78-1.26] | 0.923 | 0.98 [0.77-1.25] | 0.861 |
|  | Cases ≤4 months | 1.22 [0.98-1.53] | 0.079 | 1.09 [0.86-1.38] | 0.492 | 1.21 [0.96-1.54] | 0.105 | 1.22 [0.96-1.55] | 0.112 |
| Age group | 18-35 years | 1 |  | 1 |  | 1 |  | 1 |  |
|  | 36-65 years | 0.43 [0.37-0.50] | <0.001 | 0.37 [0.32-0.44] | <0.001 | 2.09 [1.81-2.41] | <0.001 | 0.49 [0.42-0.58] | <0.001 |
|  | 65+ years | 0.32 [0.27-0.39] | <0.001 | 0.20 [0.17-0.24] | <0.001 | 3.60 [3.00-4.34] | <0.001 | 0.23 [0.19-0.28] | <0.001 |
| Educational level | High | 1 |  | 1 |  | 1 |  | 1 |  |
|  | Intermediate | 1.01 [0.89-1.14] | 0.904 | 0.85 [0.74-0.96] | 0.013 | 1.55 [1.36-1.77] | <0.001 | 0.96 [0.84-1.09] | 0.510 |
|  | Low | 0.98 [0.84-1.15] | 0.833 | 0.77 [0.66-0.90] | 0.001 | 1.86 [1.58-2.21] | <0.001 | 0.89 [0.76-1.04] | 0.139 |
| Sex | Male | 1 |  | 1 |  | 1 |  | 1 |  |
|  | Female | 1.47 [1.31-1.65] | <0.001 | 1.59 [1.42-1.79] | <0.001 | 1.17 [1.03-1.31] | 0.012 | 1.60 [1.42-1.79] | <0.001 |
| Comorbidities | No comorbidities | 1 |  | 1 |  | 1 |  | 1 |  |
|  | 1 comorbidity | 1.46 [1.27-1.67] | <0.001 | 1.10 [0.96-1.27] | 0.162 | 1.92 [1.67-2.22] | <0.001 | 1.50 [1.30-1.73] | <0.001 |
|  | 2+ comorbidities | 1.95 [1.59-2.39] | <0.001 | 1.16 [0.94-1.42] | 0.163 | 3.51 [2.71-4.60] | <0.001 | 2.94 [2.35-3.70] | <0.001 |
| Stringency index | 75 | 1 |  | 1 |  | 1 |  | 1 |  |
|  | 79 | 0.97 [0.90-1.05] | 0.457 | 0.96 [0.89-1.04] | 0.381 | 0.98 [0.90-1.06] | 0.576 | 0.91 [0.84-0.99] | 0.030 |

Table 17. Results of the four multivariable logistic regression models for health utility (SF-6D), mental health (MCS), physical health (PCS) and fatigue (CIS) at the 65% cut-off of the cumulative distribution of uninfected.

|  |  | SF-6D | | MCS | | PCS | | CIS | |
| --- | --- | --- | --- | --- | --- | --- | --- | --- | --- |
|  |  | **OR [CI]** | **p-value** | **OR [CI]** | **p-value** | **OR [CI]** | **p-value** | **OR [CI]** | **p-value** |
|  | Intercept | 10.99 | 0.428 | 8.20 | 0.530 | 1.70 | 0.867 | 2274.98 | 0.025 |
| Serological status | Uninfected | 1 |  | 1 |  | 1 |  | 1 |  |
|  | Cases >4 months | 1.05 [0.83-1.32] | 0.702 | 0.93 [0.73-1.20] | 0.591 | 1.01 [0.79-1.29] | 0.950 | 0.94 [0.74-1.21] | 0.645 |
|  | Cases ≤4 months | 1.22 [0.98-1.53] | 0.079 | 1.00 [0.78-1.28] | 0.998 | 1.12 [0.88-1.42] | 0.370 | 1.19 [0.93-1.53] | 0.165 |
| Age group | 18-35 years | 1 |  | 1 |  | 1 |  | 1 |  |
|  | 36-65 years | 0.43 [0.37-0.50] | <0.001 | 0.36 [0.30-0.43] | <0.001 | 1.90 [1.64-2.19] | <0.001 | 0.51 [0.43-0.60] | <0.001 |
|  | 65+ years | 0.32 [0.27-0.39] | <0.001 | 0.18 [0.15-0.22] | <0.001 | 3.55 [2.94-4.31] | <0.001 | 0.24 [0.20-0.30] | <0.001 |
| Educational level | High | 1 |  | 1 |  | 1 |  | 1 |  |
|  | Intermediate | 1.01 [0.89-1.14] | 0.904 | 0.77 [0.67-0.88] | <0.001 | 1.62 [1.41-1.85] | <0.001 | 0.94 [0.82-1.07] | 0.355 |
|  | Low | 0.98 [0.84-1.15] | 0.833 | 0.71 [0.61-0.83] | <0.001 | 1.88 [1.58-2.24] | <0.001 | 0.88 [0.75-1.04] | 0.124 |
| Sex | Male | 1 |  | 1 |  | 1 |  | 1 |  |
|  | Female | 1.47 [1.31-1.65] | <0.001 | 1.63 [1.45-1.84] | <0.001 | 1.15 [1.02-1.30] | 0.024 | 1.62 [1.43-1.82] | <0.001 |
| Comorbidities | No comorbidities | 1 |  | 1 |  | 1 |  | 1 |  |
|  | 1 comorbidity | 1.46 [1.27-1.67] | <0.001 | 0.98 [0.85-1.13] | 0.746 | 1.86 [1.60-2.16] | <0.001 | 1.47 [1.27-1.70] | <0.001 |
|  | 2+ comorbidities | 1.95 [1.59-2.39] | <0.001 | 0.97 [0.79-1.19] | 0.779 | 3.06 [2.34-4.07] | <0.001 | 2.80 [2.23-3.54] | <0.001 |
| Stringency index | 75 | 1 |  | 1 |  | 1 |  | 1 |  |
|  | 79 | 0.97 [0.90-1.05] | 0.457 | 0.99 [0.91-1.08] | 0.871 | 0.99 [0.91-1.07] | 0.725 | 0.92 [0.84-1.00] | 0.050 |

Table 18. Results of the four multivariable logistic regression models for health utility (SF-6D), mental health (MCS), physical health (PCS) and fatigue (CIS) at the 70% cut-off of the cumulative distribution of uninfected.

|  |  | SF-6D | | MCS | | PCS | | CIS | |
| --- | --- | --- | --- | --- | --- | --- | --- | --- | --- |
|  |  | **OR [CI]** | **p-value** | **OR [CI]** | **p-value** | **OR [CI]** | **p-value** | **OR [CI]** | **p-value** |
|  | Intercept | 10.99 | 0.428 | 73.54 | 0.228 | 0.42 | 0.785 | 15876.00 | 0.012 |
| Serological status | Uninfected | 1 |  | 1 |  | 1 |  | 1 |  |
|  | Cases >4 months | 1.05 [0.83-1.32] | 0.702 | 0.85 [0.66-1.10] | 0.210 | 0.97 [0.76-1.26] | 0.842 | 0.98 [0.76-1.28] | 0.886 |
|  | Cases ≤4 months | 1.22 [0.98-1.53] | 0.079 | 1.01 [0.79-1.31] | 0.917 | 1.00 [0.78-1.28] | 0.991 | 1.25 [0.96-1.65] | 0.103 |
| Age group | 18-35 years | 1 |  | 1 |  | 1 |  | 1 |  |
|  | 36-65 years | 0.43 [0.37-0.50] | <0.001 | 0.37 [0.30-0.45] | <0.001 | 1.97 [1.71-2.28] | <0.001 | 0.49 [0.40-0.58] | <0.001 |
|  | 65+ years | 0.32 [0.27-0.39] | <0.001 | 0.18 [0.15-0.23] | <0.001 | 3.76 [3.08-4.60] | <0.001 | 0.23 [0.19-0.28] | <0.001 |
| Educational level | High | 1 |  | 1 |  | 1 |  | 1 |  |
|  | Intermediate | 1.01 [0.89-1.14] | 0.904 | 0.80 [0.70-0.92] | 0.002 | 1.59 [1.38-1.83] | <0.001 | 0.96 [0.83-1.11] | 0.556 |
|  | Low | 0.98 [0.84-1.15] | 0.833 | 0.73 [0.62-0.86] | <0.001 | 2.00 [1.66-2.41] | <0.001 | 0.81 [0.69-0.96] | 0.013 |
| Sex | Male | 1 |  | 1 |  | 1 |  | 1 |  |
|  | Female | 1.47 [1.31-1.65] | <0.001 | 1.61 [1.42-1.83] | <0.001 | 1.16 [1.03-1.32] | 0.018 | 1.60 [1.41-1.82] | <0.001 |
| Comorbidities | No comorbidities | 1 |  | 1 |  | 1 |  | 1 |  |
|  | 1 comorbidity | 1.46 [1.27-1.67] | <0.001 | 1.05 [0.91-1.22] | 0.514 | 1.73 [1.48-2.02] | <0.001 | 1.42 [1.22-1.66] | <0.001 |
|  | 2+ comorbidities | 1.95 [1.59-2.39] | <0.001 | 1.13 [0.91-1.40] | 0.271 | 2.91 [2.18-3.95] | <0.001 | 2.68 [2.10-3.45] | <0.001 |
| Stringency index | 75 | 1 |  | 1 |  | 1 |  | 1 |  |
|  | 79 | 0.97 [0.90-1.05] | 0.457 | 0.97 [0.88-1.06] | 0.473 | 1.01 [0.93-1.09] | 0.875 | 0.90 [0.81-0.99] | 0.033 |

Table 19. Results of the four multivariable logistic regression models for health utility (SF-6D), mental health (MCS), physical health (PCS) and fatigue (CIS) at the 75% cut-off of the cumulative distribution of uninfected.

|  |  | SF-6D | | MCS | | PCS | | CIS | |
| --- | --- | --- | --- | --- | --- | --- | --- | --- | --- |
|  |  | **OR [CI]** | **p-value** | **OR [CI]** | **p-value** | **OR [CI]** | **p-value** | **OR [CI]** | **p-value** |
|  | Intercept | 10.99 | 0.428 | 81.06 | 0.250 | 0.10 | 0.483 | 469.71 | 0.116 |
| Serological status | Uninfected | 1 |  | 1 |  | 1 |  | 1 |  |
|  | Cases >4 months | 1.05 [0.83-1.32] | 0.702 | 1.00 [0.76-1.33] | 0.985 | 0.98 [0.76-1.28] | 0.882 | 0.93 [0.71-1.22] | 0.582 |
|  | Cases ≤4 months | 1.22 [0.98-1.53] | 0.079 | 0.98 [0.75-1.30] | 0.909 | 1.00 [0.78-1.29] | 0.988 | 1.22 [0.92-1.63] | 0.176 |
| Age group | 18-35 years | 1 |  | 1 |  | 1 |  | 1 |  |
|  | 36-65 years | 0.43 [0.37-0.50] | <0.001 | 0.33 [0.26-0.40] | <0.001 | 2.02 [1.74-2.34] | <0.001 | 0.48 [0.39-0.58] | <0.001 |
|  | 65+ years | 0.32 [0.27-0.39] | <0.001 | 0.15 [0.12-0.19] | <0.001 | 3.70 [3.00-4.57] | <0.001 | 0.21 [0.17-0.27] | <0.001 |
| Educational level | High | 1 |  | 1 |  | 1 |  | 1 |  |
|  | Intermediate | 1.01 [0.89-1.14] | 0.904 | 0.78 [0.67-0.91] | 0.001 | 1.53 [1.33-1.77] | <0.001 | 0.94 [0.81-1.10] | 0.435 |
|  | Low | 0.98 [0.84-1.15] | 0.833 | 0.74 [0.63-0.88] | 0.001 | 2.01 [1.65-2.46] | <0.001 | 0.79 [0.67-0.94] | 0.008 |
| Sex | Male | 1 |  | 1 |  | 1 |  | 1 |  |
|  | Female | 1.47 [1.31-1.65] | <0.001 | 1.71 [1.50-1.95] | <0.001 | 1.15 [1.01-1.31] | 0.033 | 1.59 [1.39-1.82] | <0.001 |
| Comorbidities | No comorbidities | 1 |  | 1 |  | 1 |  | 1 |  |
|  | 1 comorbidity | 1.46 [1.27-1.67] | <0.001 | 1.05 [0.90-1.23] | 0.535 | 1.74 [1.48-2.06] | <0.001 | 1.44 [1.23-1.69] | <0.001 |
|  | 2+ comorbidities | 1.95 [1.59-2.39] | <0.001 | 1.13 [0.91-1.41] | 0.274 | 3.01 [2.19-4.23] | <0.001 | 2.79 [2.16-3.65] | <0.001 |
| Stringency index | 75 | 1 |  | 1 |  | 1 |  | 1 |  |
|  | 79 | 0.97 [0.90-1.05] | 0.457 | 0.97 [0.88-1.07] | 0.548 | 1.03 [0.95-1.11] | 0.511 | 0.94 [0.85-1.04] | 0.250 |

Figure 1. Post-hoc analysis of health utility scores (SF6-D) portraying the minimal difference in prevalence between the cases >4 months and those uninfected for significance in the logistic regression model at each cut-off point.

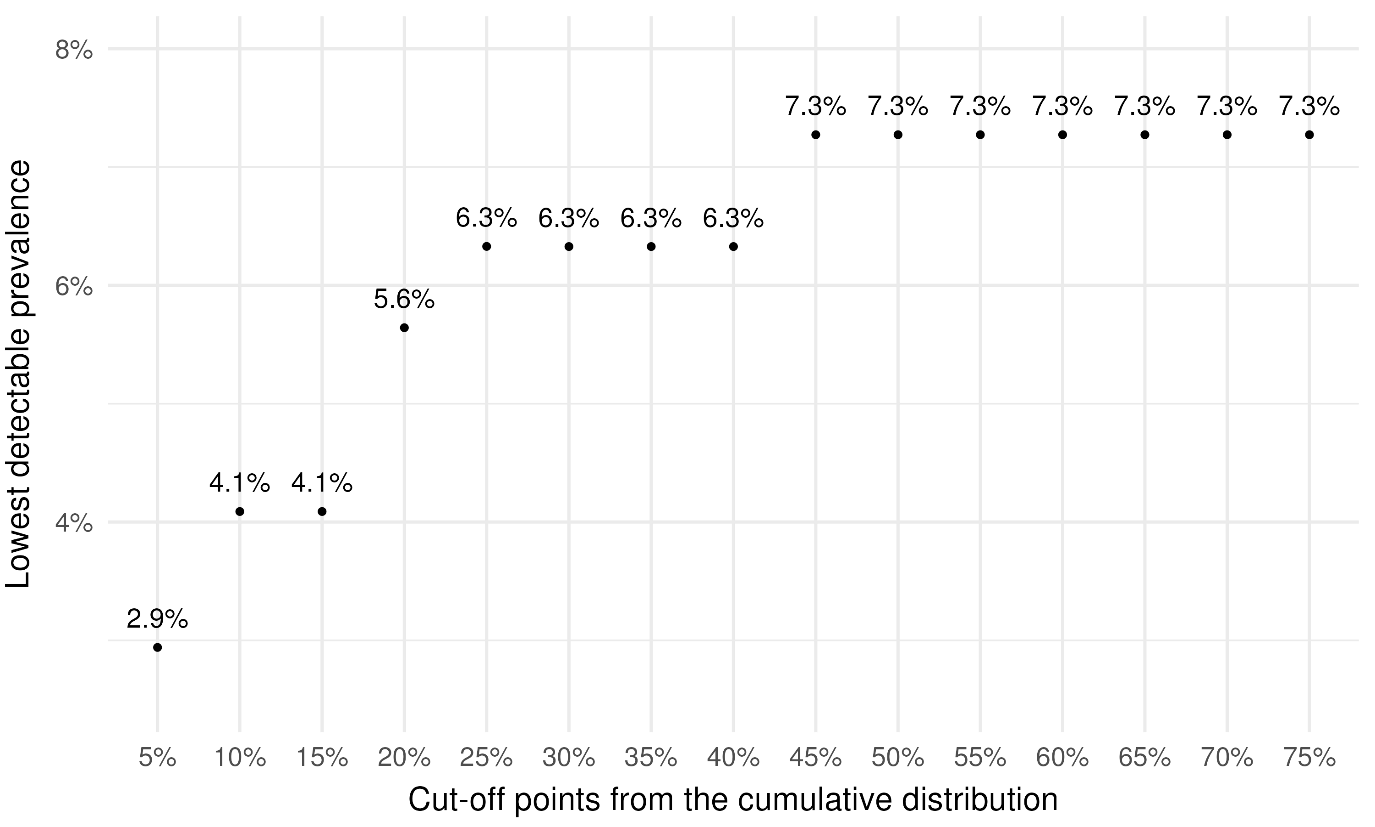

Figure 2. Post-hoc analysis of mental health scores (MCS) portraying the minimal difference in prevalence between the cases >4 months and those uninfected for significance in the logistic regression model at each cut-off point.

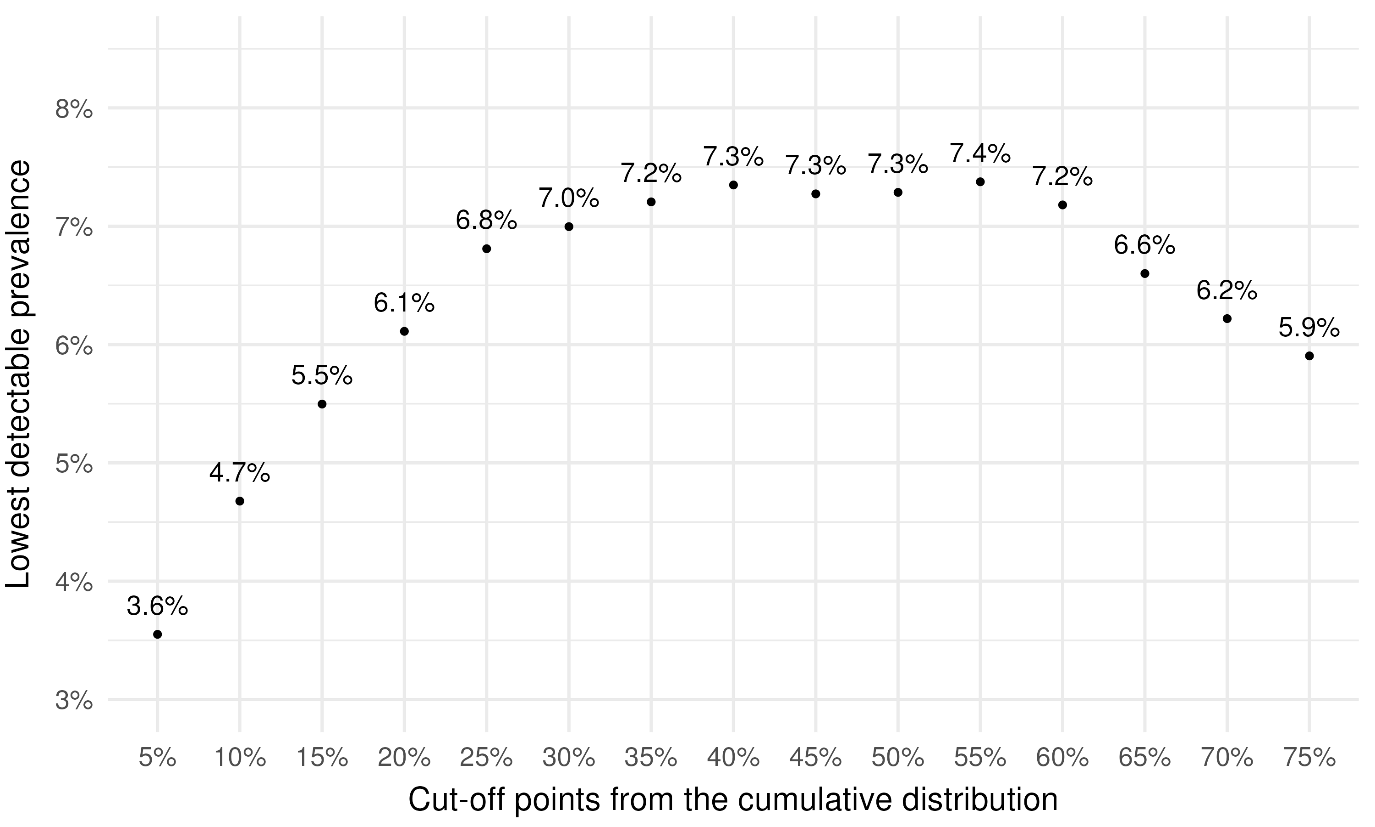

Figure 3. Post-hoc analysis of physical health scores (PCS) portraying the minimal difference in prevalence between the cases >4 months and those uninfected for significance in the logistic regression model at each cut-off point.

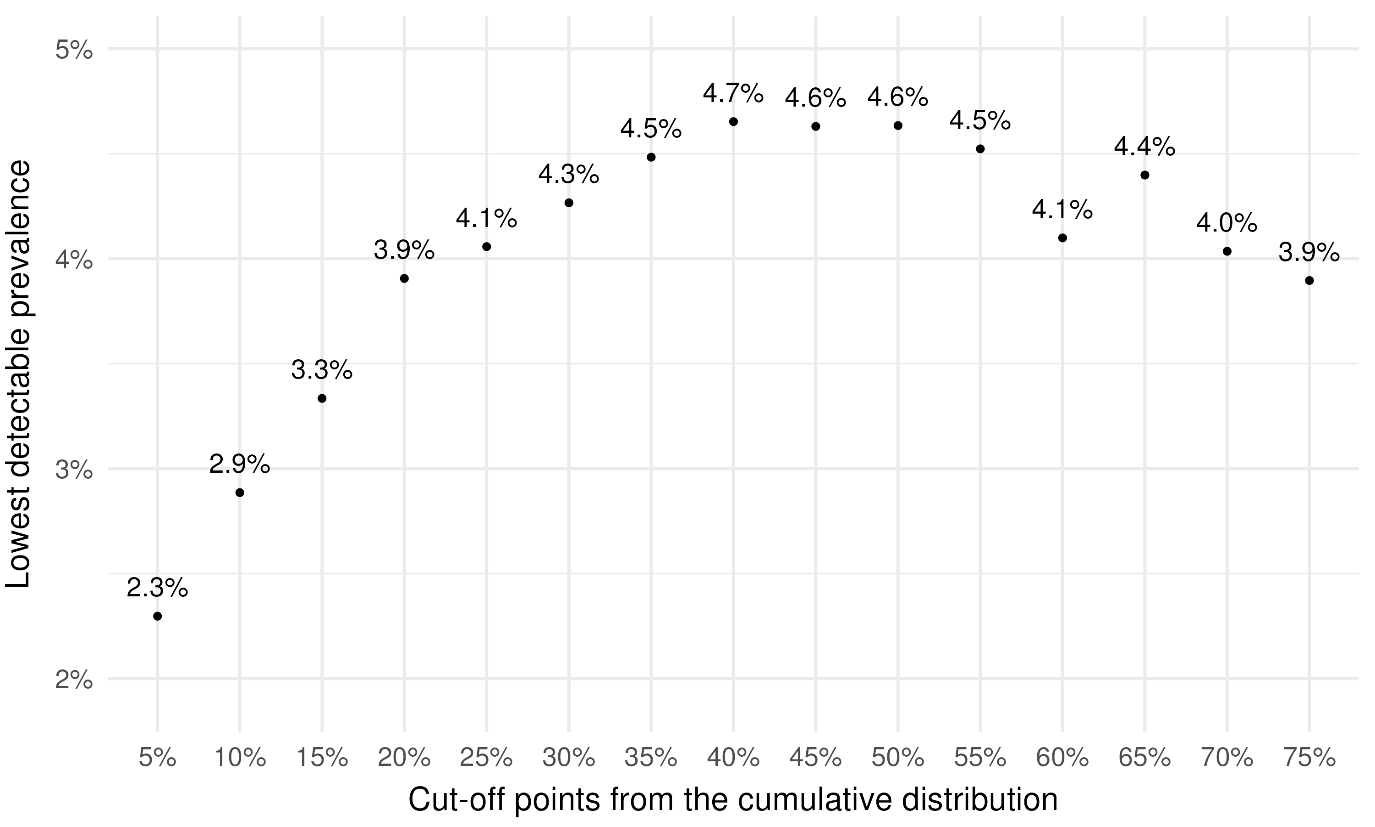

Figure 4. Post-hoc analysis of fatigue scores (CIS) portraying the minimal difference in prevalence between the cases >4 months and those uninfected for significance in the logistic regression model at each cut-off point.

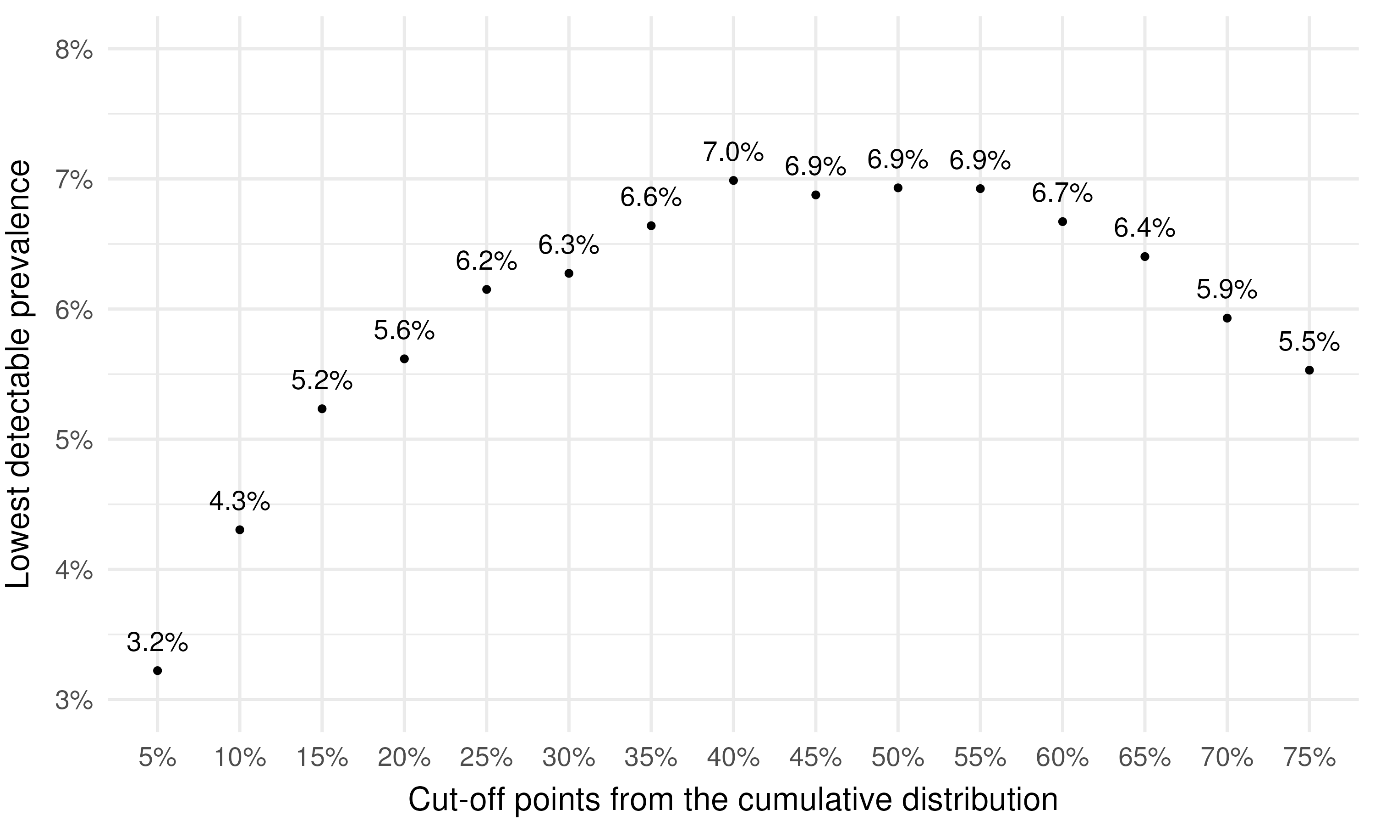
